# Steroid hormone levels vary with sex, aging, lifestyle, and genetics

**DOI:** 10.1101/2024.10.07.24315000

**Authors:** Léa G Deltourbe, Jamie Sugrue, Elizabeth Maloney, Florian Dubois, Anthony Jaquaniello, Jacob Bergstedt, Etienne Patin, Lluis Quintana-Murci, Molly A Ingersoll, Darragh Duffy, Milieu Intérieur Consortium

## Abstract

Steroid hormone levels vary greatly among individuals, between the sexes, with age, and across health and disease states. Nevertheless, what drives variance in steroid hormones and, globally, how steroid hormones vary in an individual over time are not well-studied. To address this fundamental gap in knowledge, we measured the levels of 17 steroid hormones in a sex-balanced cohort of 949 healthy donors ranging in age from 20-69 years. We investigated associations between steroid levels and biological sex, age, clinical and demographic data, genetics, and proteomics. Steroid hormone levels were strongly affected by biological sex and age as hypothesized, but also associated with a surprisingly high number of lifestyle habits. Key among our observations was the broad impact of hormonal birth control in female donors and the relationship with smoking in male donors. Using data collected from a 10-year follow-up of the cohort, we identified significant associations between steroid hormone levels and health status only in male donors. These observations provide a unique and comprehensive resource for steroid hormone level variance in healthy individuals and highlight biological and lifestyle parameters that can impact these levels. Our findings underly the importance of considering parameters, such as sex, age, and potentially gendered behaviors, in preventative health care and the treatment of hormone-related diseases.

**One sentence summary:** **Circulating steroid hormone levels are influenced by biological sex, age, lifestyle behaviors, and host genetics.**

## Introduction

Steroid hormones are key biomolecules regulating diverse and complex developmental and physiological processes^1–3^. Steroid hormones can be separated into two groups, and further divided into five sub-groups. The first group, the corticoids, includes mineralocorticoids and glucocorticoids and the second group, the sex hormones, is composed of progestogens, estrogens, and androgens. Their functions include cell metabolism, immune cell behavior, resistance to stress, water regulation and salt balance, and development and maintenance of the reproductive system and secondary sexual characteristics. Steroid hormone levels can vary greatly between the biological sexes and with age. This variability plays an underestimated critical role in immune responses and disease risk and outcome^4–6^.

Sex, age, and reproductive cycle changes in steroid hormone levels are involved in many sex- and age-associated pathophysiologies and immune responses^7,8^. For example, elevated cortisol causes Cushing’s syndrome, while Addison’s disease is a result of adrenal insufficiency and low cortisol levels^9,10^. Both conditions are more prevalent in women and peak incidence occurs between 30-50 years of age^9,10^. Sex steroids, such as testosterone, impact asthma differentially in women and men with age, in that young boys are more susceptible than girls to this disease, but after puberty, women have higher incidence^11^. In certain pathologies, such as polycystic ovary syndrome, in which women have atypically high androgen levels, it is unclear whether the disease is the cause or the result of steroid hormone dysregulation^7,12^. Overall, dysregulation in steroid expression levels can impact the development of many diseases such as rheumatoid arthritis or reproductive cancers, while others, such as sepsis or HIV can cause steroid expression level dysregulation^7,8,13,14^. Given their ability to induce or inhibit inflammation, steroid hormones (in particular corticosteroids) are major therapies used in rheumatic diseases^15^, asthma^16^ and even infection^17^. Direct inhibition of sex steroids is also used as a therapy to treat malignancies such as breast or prostate cancer^18^. Overall, steroid levels are good indicators of health status, therefore, defining healthy ranges and understanding how they change over time may support new preventative and therapeutic approaches for a wide range of diseases.

Despite their clear role in homeostasis and immunity, studies of intra- and interindividual differences in steroid levels between females and males or across ages are strikingly limited. Many studies focus on one or a few steroids, without consideration of the impact of upstream or intermediate molecules and interconnected steroidogenesis pathways, or use techniques poorly adapted to measuring these molecules^19^. To address this large gap in our knowledge, we measured the levels of 17 steroid hormones by targeted mass spectrometry in the *Milieu Intérieur* cohort of 949 healthy donors stratified for sex and age (half female, half male from 20-69 years of ages)^20^. We found that nearly all steroids exhibited sex or age-related variation, hormonal birth control profoundly repressed the estrogens, progestogens, and androgens to levels lower than those found in menopausal women, and testosterone decreases may be specifically associated with declining health status, and not only with aging.

## Results

### Steroid classes cluster distinctly between the sexes

To determine interindividual variation in a healthy human cohort, we measured the levels of 17 major steroid hormones, with representation of the five subgroups: 11-deoxycorticosterone, corticosterone, aldosterone (mineralocorticoids); 11-deoxycortisol, cortisol, cortisone (glucocorticoids); progesterone, 17α-hydroxyprogesterone (17-OHP) (progestogens); estrone (E1), estradiol (E2) (estrogens); dehydroepiandrosterone (DHEA), dehydroepiandrosterone sulfate (DHEAS), etiocholanolone, androstenedione, androsterone, testosterone, and dihydrotestosterone (DHT) (androgens), in the plasma of 1,000 donors by liquid chromatography coupled with mass spectrometry (LC-MS/MS). 51 donors were excluded from downstream analysis either because their sample could not be processed due to viscosity (7 donors) or they did not consent to make their data publicly available (44 donors), resulting in a final sample size of 949. Donors self-declared their gender in the initial recruitment visit (V0)^21^. To specifically study biological sex as a variable and identify differences between female and male donors, we relied upon allosome (X and Y chromosomes) karyotype, identified by whole genome SNP arrays^21^.

Applying principal component analysis (PCA) to reduce data dimensionality, we observed that steroid levels broadly clustered by group, in which the mineralocorticoids and glucocorticoids were closely clustered with each other (**Figure 1A**). Progesterone and the estrogens clustered together, as did the androgens DHT and testosterone. The remaining androgens and the progestogen 17-OHP clustered together, distinct from the rest of the steroids. Pairwise correlation analysis showed that those that clustered the most closely together were the most correlated steroids, such as the mineralocorticoid corticosterone and glucocorticoid cortisol (**Figure 1B**). The androgens DHEA, DHEAS, androstenedione, and androsterone had strong positive correlations amongst each other and with 17-OHP. Given that testosterone is a precursor for DHT, and E1 and E2 can be converted into each other, their strong positive correlations were expected (**Fig S1, S2**). Sex specific correlation analysis showed differences in the strengths of the steroid relationships between males and females (**Fig S3**). For instance, DHT correlated with both DHEA and DHEAS in females (r2 = 0.53 and 0.49 respectively) but not in males (r^2 = 0.06 and -0.01) (**Fig S3**).

**Figure 1.**
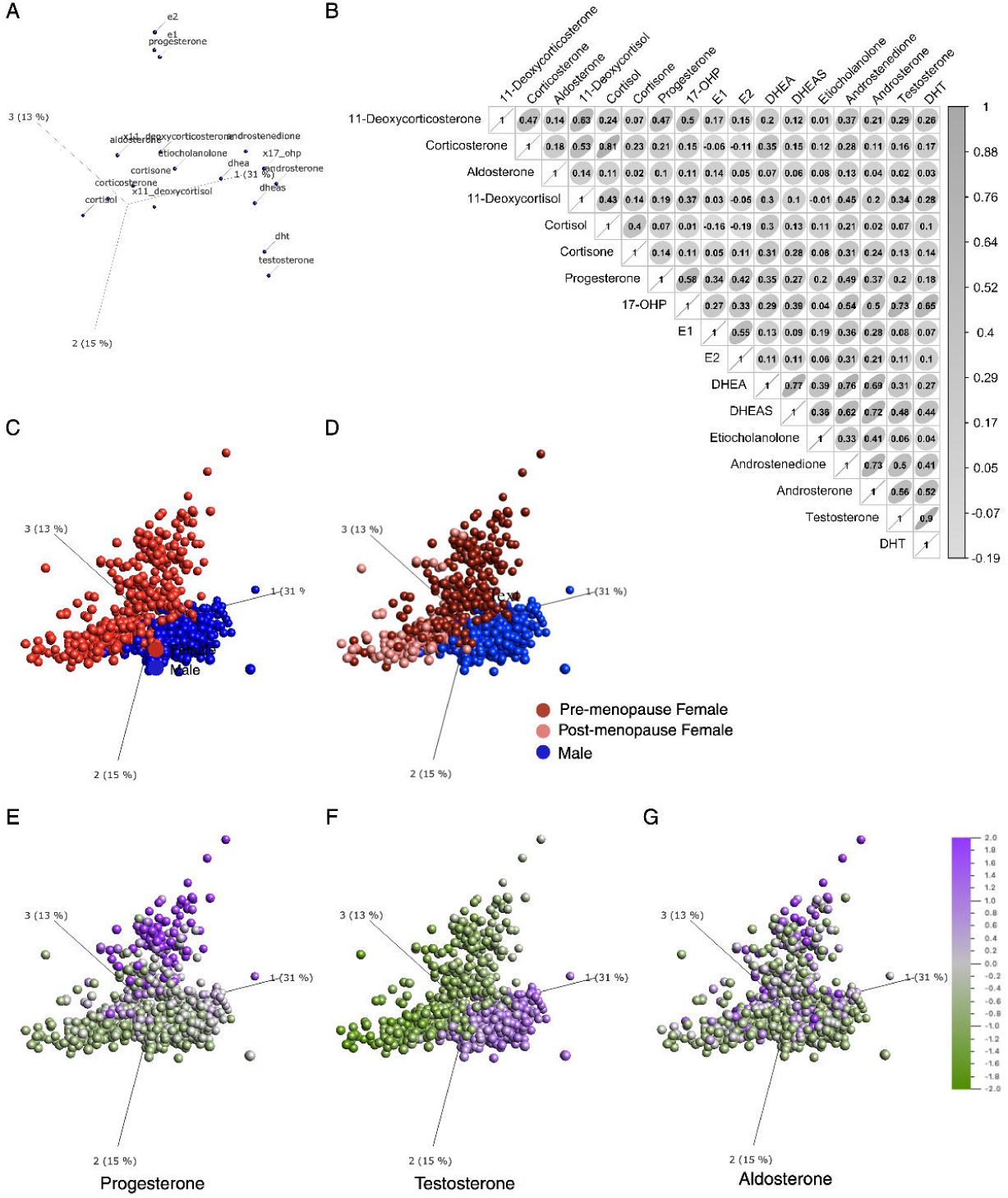
Steroid hormones and donors cluster by group and sex. We applied PCA to the log_2_-transformed nanomolar concentrations of steroid hormones measured by LC-MS/MS across 949 donors. (A) PCA shows clustering of steroid hormones across the first three principal components. (B) Correlation matrix showing Spearman correlations between hormones for all donors. Ellipses indicate the direction of correlation. (C-D) PCA shows individual donors color-coded by (C) sex or (D) sex and menopausal status in female donors. (E-G) Expression levels of (E) progesterone, (F) testosterone, and (G) aldosterone overlaid on the PCA plot. (A, C-G) Proportion of variance explained by PC1, PC2, and PC3 are reported on the axes. (n=949 donors, 17 variables).

Principal components 1, 2 and 3, which together explain 59% of variance in steroid hormone levels, were strongly aligned with sex (**Figure 1C or 1D**). Progesterone and estrogen levels and DHT and testosterone levels clearly drove sex-specific clustering of the donors (**Figure 1C**), which could be further resolved by distinguishing pre-and post-menopausal women in the colored overlays (**Figure 1D**). No donor was perimenopausal at the time of recruitment^22^. Notably, sex steroids, such as progesterone (**Figure 1E**) and testosterone (**Figure 1F**) displayed the starkest differences between the sexes, whereas the mineralocorticoid aldosterone (**Figure 1G**) had variable expression levels between the sexes and among pre- and post-menopausal women. The two corticoid groups had expression levels that did not strongly associate with female or male donors and the remaining sex hormones showed the expected sex-specific patterns (**Fig S3)**.

### Age impacts steroid levels differently in female and male donors

To describe variability in individual steroid levels as a function of age and sex, we plotted log_2_-transformed nanomolar concentrations and applied locally estimated scatterplot smoothing (LOESS) models on the data (**Figure 2A**). To determine the strength of the relationships between steroid hormone levels and sex or age, we applied linear models to the log_2_-transformed nanomolar concentrations and calculated effect sizes on steroid levels, for age separately among female or male donors (**Figure 2B, C)**, for sex **(Figure 2D, E)**, and for age×sex interactions **(Figure 2F)**. To aid in interpreting the age×sex interactions, the effect sizes from the models applied with female donors as a reference (**Figure 2B, D)** and male donors as a reference are shown (**Figure 2C, E).** Strikingly, 12 out of the 17 steroid hormones had a significant age×sex interaction (FDR *q*-value<0.05) (**Figure 2F**). Among the mineralocorticoids, 11-deoxycorticosterone did not change with age or between the sexes, whereas corticosterone and aldosterone levels declined with age, which was more pronounced in female donors (**Figure 2A-C**) as illustrated by the age×sex interaction (**Figure 2F**). Levels of the glucocorticoids 11-deoxycortisol and cortisol were significantly different between the sexes. These differences were more pronounced in younger donors with a significant age effect in female donors only and an age×sex interaction effect on cortisol (**Figure 2A, B, D-F)**. Interestingly, age was negatively correlated with cortisol and positively correlated with its precursor 11-deoxycortisol **(Figure 2B**), suggesting age-related changes in cortisol biosynthesis. Cortisone levels were similar between female and male donors and declined with age, which was more pronounced in female donors as reflected in the significant age×sex interaction (**Figure 2A-C, F**).

**Figure 2.**
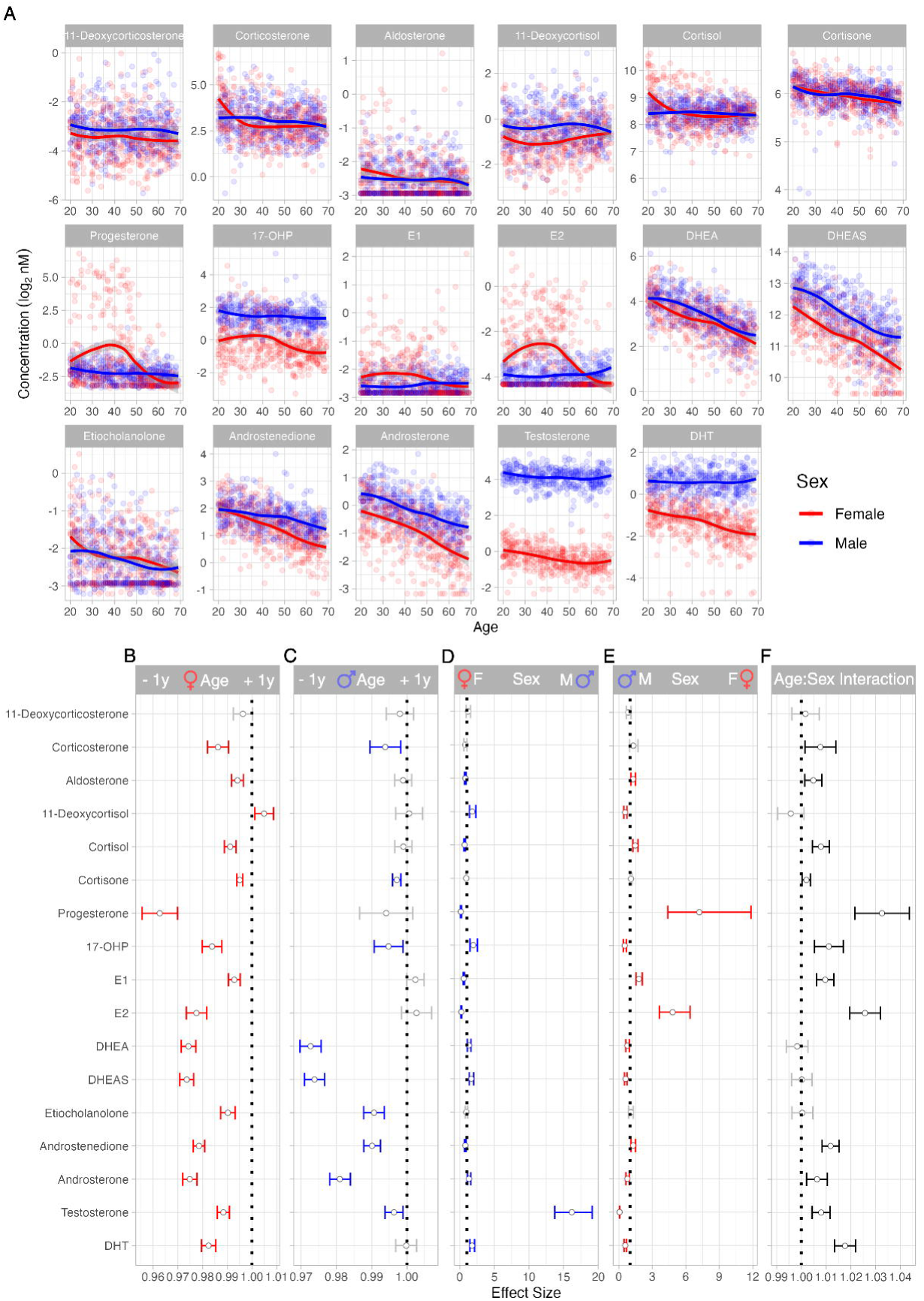
Steroid hormone levels differ with age and sex. (**A**) Scatter plots show steroid hormone nanomolar log_2_ transformed concentrations by age with applied LOESS models. Female donors are indicated in red, male donors in blue. (**B-F**) Linear models were applied to quantify the effects of (**B**) age in female donors, (**C**) age in male donors, (**D**) sex with female donors as the reference, (**E**) sex with male donors as the reference, and (**F**) the interaction of age and sex on steroid hormone levels. FDR correction for multiple testing was applied globally for all 3 terms (age, sex, age×sex interaction) over 17 steroid hormone variables. Significant effects (FDR q-value <0.05) are indicated in red (female), blue (male), or black (both sexes), while non-significant effects are in grey. (n=949 donors).

From the mid-40s, progesterone levels dropped precipitously in female donors reflecting the proportional increase of menopausal donors from this age. By contrast, progesterone levels in male donors were low and relatively stable over time (**Figure 2A-C**). 17-OHP levels were significantly higher in male donors compared to female donors (**Figure 2A, D, E**) and gradually declined with age in both sexes (**Figure 2A-C**). Estrogens E1 and E2 were higher in women and declined with age, whereas these hormones were less variable over time in men (**Figure 2**). Finally, the androgens, DHEA, DHEAS, androstenedione, androsterone, testosterone, and DHT were higher in male donors compared to female donors (**Figure 2D, E**). Etiocholanolone was not significantly different between female and male donors, potentially due to a high number of samples being at the limit of detection (**Figure 2A, D, E**). All the androgens measured significantly declined with age for both sexes, with the surprising exception of DHT, which did not decline in male donors, as reflected in the significant age×sex interaction (**Figure 2A-C, F**). We observed that testosterone levels were reduced in older donors (**Figure 2E, F**), but not as drastically as previous reports of testosterone decline in aging men, the so-called andropause^7^.

### Lifestyle behaviors are markedly associated with steroid level differences

We next determined whether specific lifestyle behaviors were associated with steroid levels, while accounting for the expected influence of age and sex. We performed likelihood ratio tests (LRT) for significant associations between the levels of each of the 17 steroids and 137 variables extracted from our study case report form (CRF), with all 949 donors. Due to the significant effect sizes found in the sex×age interaction analysis (**Figure 2F**), we controlled regression models for age, sex, and the interaction between sex and age. Of 137 variables tested, we identified significant associations (FDR *q*-value < 0.05) with at least one steroid for 37 variables, in categories such as physiology, diet and alcohol consumption, physical activity and sleep, education and cohabitation, smoking, and medication, among others (**Figure 3A**). BMI, weight, and abdominal circumference were associated with corticosterone and DHT levels (FDR *q*-value for BMI = 6.376e-11, for weight = 2.947e-06, and for abdominal circumference = 6.005e-08). Interestingly, DHT was also associated with a metabolic health score, which measures the cumulative risk for metabolic disease^23^. The association of physiology-based categories with steroid levels was anticipated, as, for example, adipose tissue contributes to steroidogenesis^24^.

**Figure 3.**
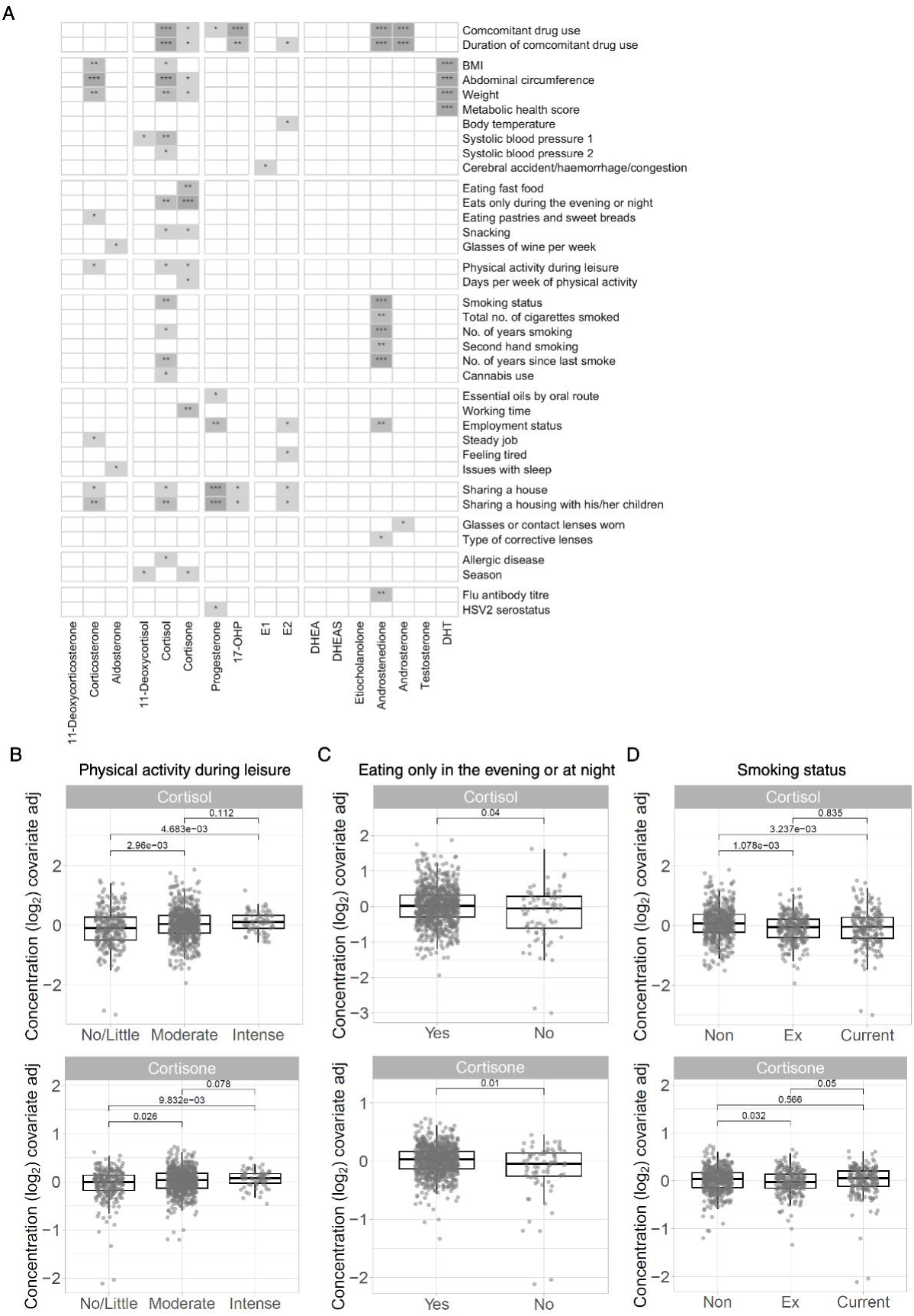
Specific characteristics and behaviors significantly associate with steroid hormone levels. (**A**) Heatmap of significant associations between steroid hormone levels and 137 CRF variables identified by likelihood ratio tests with correction for sex, age, and sex:age interactions applying FDR correction (* = *q*<0.05, * = *q*<0.01, *** = *q*<0.001). (**B-D**) Residual plots of the corrected values for cortisol and cortisone levels for (**B**) physical activity during leisure time (no/little, moderate, or intense physical activity), (**C**) eating only in evening or at night (yes, no), and (**D**) smoking status (non: non-smoker, ex: ex-smoker, current: current smoker). *q-*values were calculated using a Kruskal Wallis with FDR correction for all tests together followed by post-hoc Dunn’s test (n=949 donors).

However, we were surprised to observe associations between steroid hormone levels and specific behaviors, such as cortisone and eating habits (FDR *q*-value for eating only at night = 3.019e-05), fatigue and estradiol (FDR *q*-value = 0.03), sharing a house and progestogens (FDR *q*-value < 0.01), smoking behavior and androstenedione levels (FDR *q*-value for smoking status = 4.55e-08), and concomitant medicine use and several glucocorticoids, progestogens, estrogens, and androgens (**Figure 3A**). To understand the nature of the associations between steroid levels and lifestyle variables, we plotted the residual values of cortisol and cortisone while controlling for sex, age and the age×sex interaction, for physical activity during leisure, eating late, and smoking status (**Figure 3B-D**). Cortisol and cortisone levels were significantly lower in donors who reported no to little physical activity compared to those who reported moderate or intense physical activity (**Figure 3B**). Interestingly, cortisol and cortisone levels were not different between donors who reported moderate physical activity compared to those who reported intense physical activity (**Figure 3B**). Donors who reported frequent snacking between meals had lower levels of cortisol compared to those who never or sometimes snacked, whereas cortisone was only significantly lower in donors who reported snacking often versus never (**Figure 3C**). Non-smokers had significantly higher levels of cortisol compared to ex- and current smokers whereas cortisone levels were lower in ex versus non-smokers (**Figure 3D**).

### Lifestyle behaviors show sex- and gender-specific associations

As the likelihood ratio test analysis of the entire cohort revealed specific associations with distinct sex hormones, we assessed whether specific lifestyle behaviors correlated with steroid levels differently between the sexes. As we previously did for the entire cohort, we performed likelihood ratio tests for significant associations between the levels of each of the 17 steroids and variables extracted from the CRF, in only women or men. For the analyses of the women, we included 14 additional variables related to contraception, hormonal treatment, and history of reproductive cancer in the family. The analysis was controlled for age for men and for age, menopausal status (pre- or post-menopausal), and the interaction between age and menopausal status for women (**Figure 4**).

**Figure 4.**
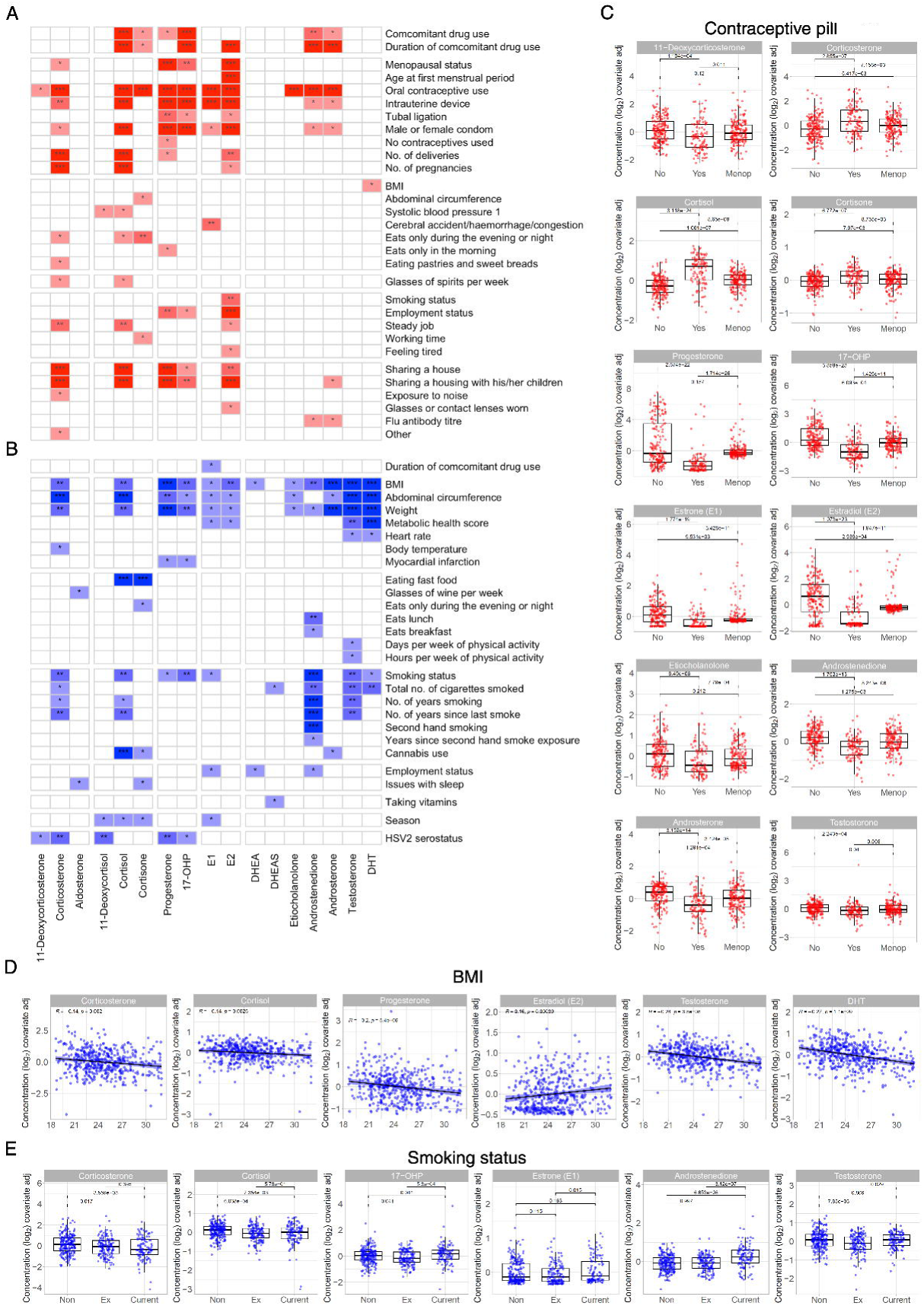
Sex-specific factors and gender-specific behaviors significantly associate with steroid hormone levels in women and men. **(A-B)** Heatmap of significant associations between steroid hormone levels and 151 or 137 CRF variables (number of variables tested in female or male donors, respectively) identified by likelihood ratio tests of **(A)** female donors with correction for age, menopausal status, and the interaction between age and menopausal status or **(B)** male donors with correction for age, applying FDR correction (* = *q*<0.05, * = *q*<0.01, *** = *q*<0.001). **(C-D)** Residual values were plotted for significant associations in **(C)** women for contraceptive pill usage (no, yes, menop = post-menopausal women) and **(D)** men for smoking status and BMI. *q-*values were calculated using a Kruskal Wallis test followed by post-hoc Dunn’s test and an FDR correction for all tests together. (n=472 (females), n=477 (males))

Of the 151 variables tested for women, we identified significant associations with at least one steroid for 30 factors, in the categories of contraception and medication, physiology, eating and working behaviors, and home ownership and living situation, among others (**Figure 4A**). Significant associations were identified between several glucocorticoid and sex steroid hormones and the additional female-specific variables we included in the analysis such as menopausal status, age of menstrual period, number of pregnancies, and type of birth control methods (**Figure 4A**). Analysis of the male donors identified 27 significantly associated variables out of the 137 tested, in categories such as physiology, eating habits and activity level, and smoking habits (**Figure 4B**). Interestingly, the significant associations we observed in the whole cohort analysis were not always present in both sexes, or did not associate with as many steroid hormones. For example, diverse eating habits or smoking variables, were significantly associated with steroid hormone levels only in men (**Figure 4A-B**). BMI, abdominal circumference, weight and metabolic score were associated with 12 different steroid levels in men, but only cortisone and DHT were significantly associated with BMI and abdominal circumference in women (**Figure 4A-B**). Conversely and surprisingly, sharing a home was only significantly associated with steroid levels in women. Whether the loss of certain significant associations is due to a loss of power (∼500 donors of each sex versus ∼1000 total donors) or these associations are driven by underlying sex or gender differences, such as those reported for smoking^25^, will require further studies.

We observed that progesterone and the estrogens declined with age, and were highly variable in premenopausal women (**Figure 2 A, B**). This variability is likely influenced by both the menstrual cycle and hormonal contraceptive pill use. Notably, hormonal contraceptive pill use was significantly associated with 12 different steroid levels, including progesterone and the estrogens (**Figure 4A**). We plotted these 12 steroid levels according to contraceptive pill use after adjusting for age, menopausal status, and the interaction between age and menopausal status (**Figure 4C**). Quite strikingly, women using oral contraceptives had significantly lower levels of 11-deoxycorticosterone, a mineralocorticoid, as well as progesterone, 17-OHP, E1, E2, and the androgens etiocholanolone, androstenedione, and androsterone compared to both women not taking contraceptive pills and post-menopausal donors (**Figure 4C**). Additionally, among pre-menopausal donors, testosterone was significantly lower in those women using hormonal contraceptive pills compared to those not using this birth control method (**Figure 4C**). Conversely, corticosterone, cortisol, and cortisone were significantly increased in women using hormonal contraceptive pills compared to donors not using contraceptive pills and post-menopausal donors (**Figure 4C**). Indeed, women not using oral hormonal birth control had median steroid hormone levels more closely aligned with post-menopausal women than women closer to their age that were using oral contraceptives, likely reflecting the inter-relatedness of steroidogenesis (**Fig S1**).

Two categories stood out for their broad associations with multiple steroid hormones in the analysis of male donors, BMI and smoking status. BMI, which in this cohort is within a healthy range (18.5-32 kg/m^2^)^22^, was significantly associated with 12 out of the 17 measured steroid hormones. We plotted residual values of 6 of the 12 most significantly associated steroid hormones (**Figure 4D**). We observed that corticosterone, cortisol, progesterone, testosterone, and DHT levels were negatively correlated with BMI (**Figure 4D**). Interestingly, estradiol levels were positively correlated with BMI. Whether the correlations are causative remains to be determined. Plotting the residuals revealed different associations between steroid levels and smoking status (**Figure 4E)**. Interestingly, corticosterone and cortisol concentrations were significantly lower in former and current smokers compared to non-smokers (**Figure 4E)**. In addition, both estrone and androstenedione concentrations were significantly higher in current smokers compared to former and non-smokers. There was no significant difference between non-smokers and current smokers, however 17-OHP and testosterone levels were lower in ex-smokers compared to the two other groups.

### Common genetic variants influence steroid hormone levels

Having found age, sex, and lifestyle factors to be associated with variation in steroid hormone levels to different extents, we next tested whether common human genetic factors influenced the 17 steroid hormone levels measured in our cohort. To do so, we performed genome wide association studies with 5,728,127 high quality variants for which the MI cohort has been genotyped^21^. To identify sex-specific genetic associations, we performed the analysis in female and male donors separately, and then performed meta-analysis on both sexes (**Figure 5A-C**). Covariates included in the model were age, BMI, smoking status, menopausal status, HRT, oral contraceptive use, IUD, and tubal ligation based on our analysis of the lifestyle data in the cohort and previous UK biobank hormone genetic association studies^26^. To account for multiple testing, a *p*-value < 5×10^-8^ was considered as significant.

**Figure 5.**
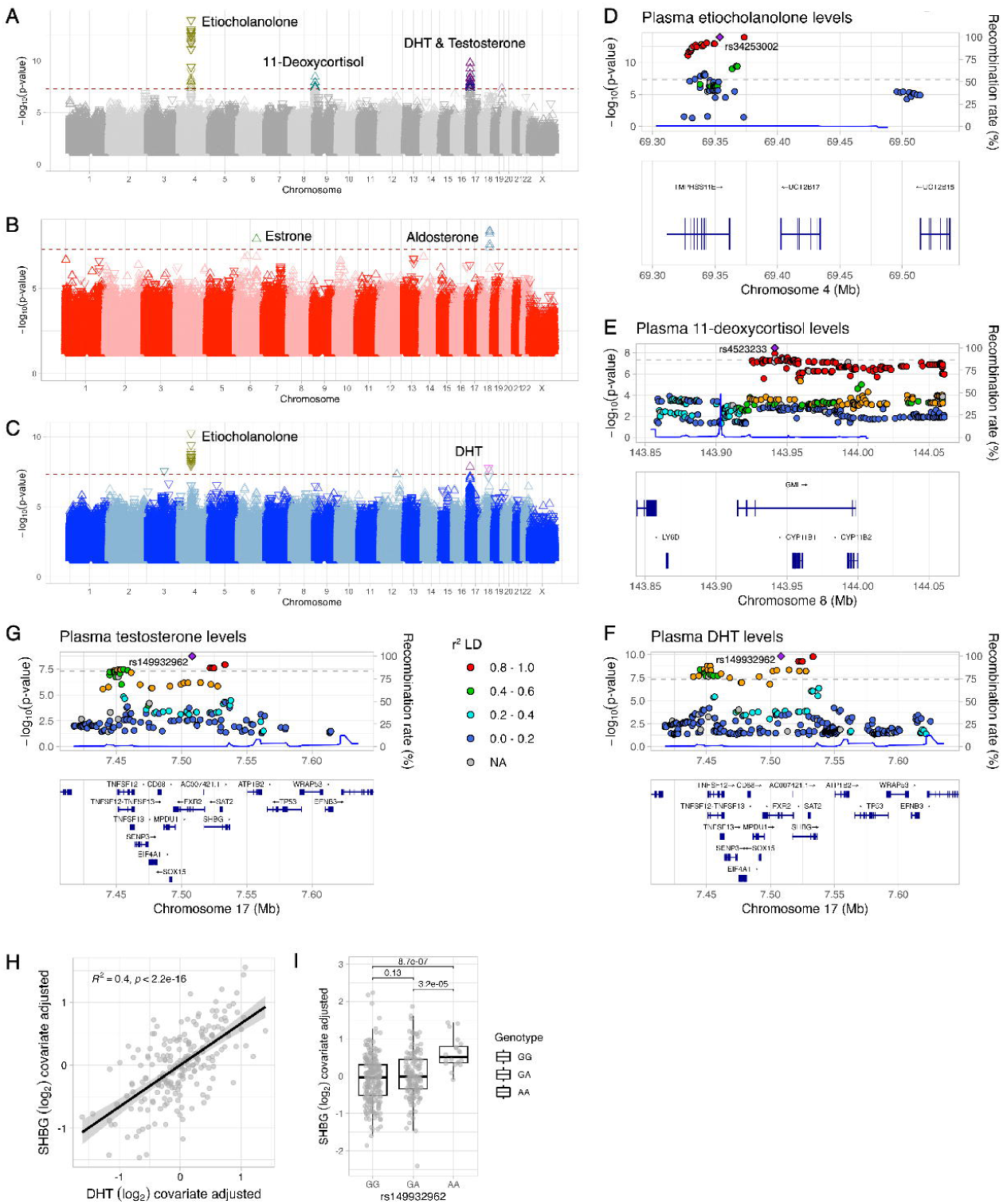
Steroid hormone levels are impacted by genetic loci. (**A-C**) Summary Manhattan plots of associations between plasma steroid hormone levels and common genetic variants in the *Milieu Intérieur* cohort for (**A**) meta-analysis of results from males and females (**B**) female donors, and (**D**) male donors. The red dotted line denotes a genome-wide significance threshold of p<5x10^-8^. The direction of the triangles indicates the effect of the variant genotype on steroid levels. (**C, E, F**) Locus zoom plots of SNPs associated with (**C**) etiocholanolone levels, (**E**) 11-deoxycortisol levels, and (**F**) DHT and (**G**) testosterone levels. The color of the locus plots denotes the level of linkage disequilibrium (r^2^) with the index SNP shown in purple. Only protein coding genes are shown in the gene tracks. The blue line indicates recombination rates. (**H**) Residual plot showing correlation between DHT and SHBG. (**I**) Residual plot showing lead testosterone and DHT variant is a pQTL for SHBG levels in a subset of *Milieu Intérieur* cohort donors (n=400). Data shown in the scatter plot (**H**) and box plot (**I**) are log_2_ data adjusted for covariates included in QTL analysis (age, BMI, smoking status, menopausal status, HRT, oral contraceptive use, IUD, tubal ligation) and by sex. Pairwise comparisons were performed on adjusted data using Kruskal-Wallis tests followed by Dunn’s postdoc test with an FDR adjustment. (n=949 donors (all), n=472 (females), n=477 (males))

In the meta-analysis of female and male results, we identified associations with three loci on chromosomes 4, 9 and 17 with etiocholanolone, 11-deoxycortisol, and DHT and testosterone respectively (**Figure 5A**). Using locus zoom plots we identified the most likely genes associated with each hormone. On chromosome 4, we identified *UGT2B17* variants associated with increased etiocholanolone levels (**Figure 5D, FigS4**). These variants are upstream of *UGT2B17*, an enzyme involved in the glucuronidation of C19 steroids such as etiocholanolone, androsterone, and testosterone^27^. *UGT2B17* is highly polymorphic and encodes one of the most commonly deleted genes in the human genome^28^. Using statistical fine mapping we identified rs34253002 to be the most likely casual variant. By exploring GTEx, an eQTL database^29^, we observed that variants such as rs2708699, which were associated with decreased levels of etiocholanolone, are associated with increased expression of *UGT2B17* (**Figure 5C**). Variants associated with an increase in etiocholanolone levels, such as rs976002, are associated with decreased expression of *UGT2B17* in whole blood and liver in GTEx^29^.

We also identified associations between 11-deoxycortisol and the *CYP11B1* locus on chromosome 8 (**Figure 5E**). CYP11B1 catalyzes the 11ϕ3-hydroxylation of 11-deoxycortisol to cortisol^30^. The SNPs identified were associated with increased levels of 11-deoxycortisol (**Fig S5**), suggesting reduced enzymatic activity or decreased expression of the enzyme. Fine mapping in a region 1Mb centered on the significant SNPs identified rs4523233 as the likely casual variant. Single cell data from GTEx indicates that the rs4523233 is associated with increased expression of CYP11B1 in the adrenal gland^29^.

Lastly, we found associations between the sex hormone binding globulin (*SHBG*) locus on chromosome 17 and testosterone and DHT levels (**Figure 5F, G, FigS6**). Fine mapping indicates that rs149932962 is the most likely casual variant associated with both DHT and testosterone. SHBG binds steroid hormones to facilitate transport of their inactive bound forms in circulation. SHBG binds DHT with high affinity and testosterone and estradiol with lower affinities. We previously measured SHBG levels in 400 individuals of the *Milieu Intérieur* cohort using Luminex multi-analyte arrays^31^. After adjusting the DHT and SHBG data for covariates included in the GWAS, we found a significant positive correlation between DHT and SHBG levels (**Figure 5H**). We also found that the lead SNP in SHBG (rs149932962) is a baseline pQTL for SHBG levels (**Figure 5I**).

In female donors specifically, we found additional associations with aldosterone levels and variants located on chromosome 18 (**Figure 5B**). These signals are located near the *TCF4* gene. In addition, a single variant near *NKAIN2* on chromosome 6 was associated with E1 levels (**Figure 5B**). In male donors, only variants that were associated with decreased etiocholanolone levels were significant. In addition, a single variant (rs149932962) in SHBG was associated with DHT levels.

### Steroid hormones are associated with plasma proteins related to physiology and inflammation

Given the previously identified associations between SHBG and DHT levels, we wanted to assess whether other plasma proteins were associated with specific steroid hormone levels. We performed correlation tests for significant associations between the levels of each of the 17 steroids and 326 soluble plasma proteins from donors aged 30 to 39 and 60 to 69 (n=371 donors in total). The analysis was first performed for all donors together for maximum statistical power (**Figure 6A**). To identify sex-specific associations, as observed in **Figure 4**, the analysis was repeated separately for male (**Figure 6B**) and female donors (**Figure 6C**). Analysis of all donors identified significant associations between 27 proteins and at least 1 steroid hormone (**Figure 6A**), many of which are found between plasma proteins and androgens in addition to progestogens which clustered together. In addition, we observed two distinct clusters, in which 10 plasma proteins were significantly positively correlated with at least one steroid hormone and 17 were significantly negatively correlated with at least one steroid hormone.

**Figure 6.**
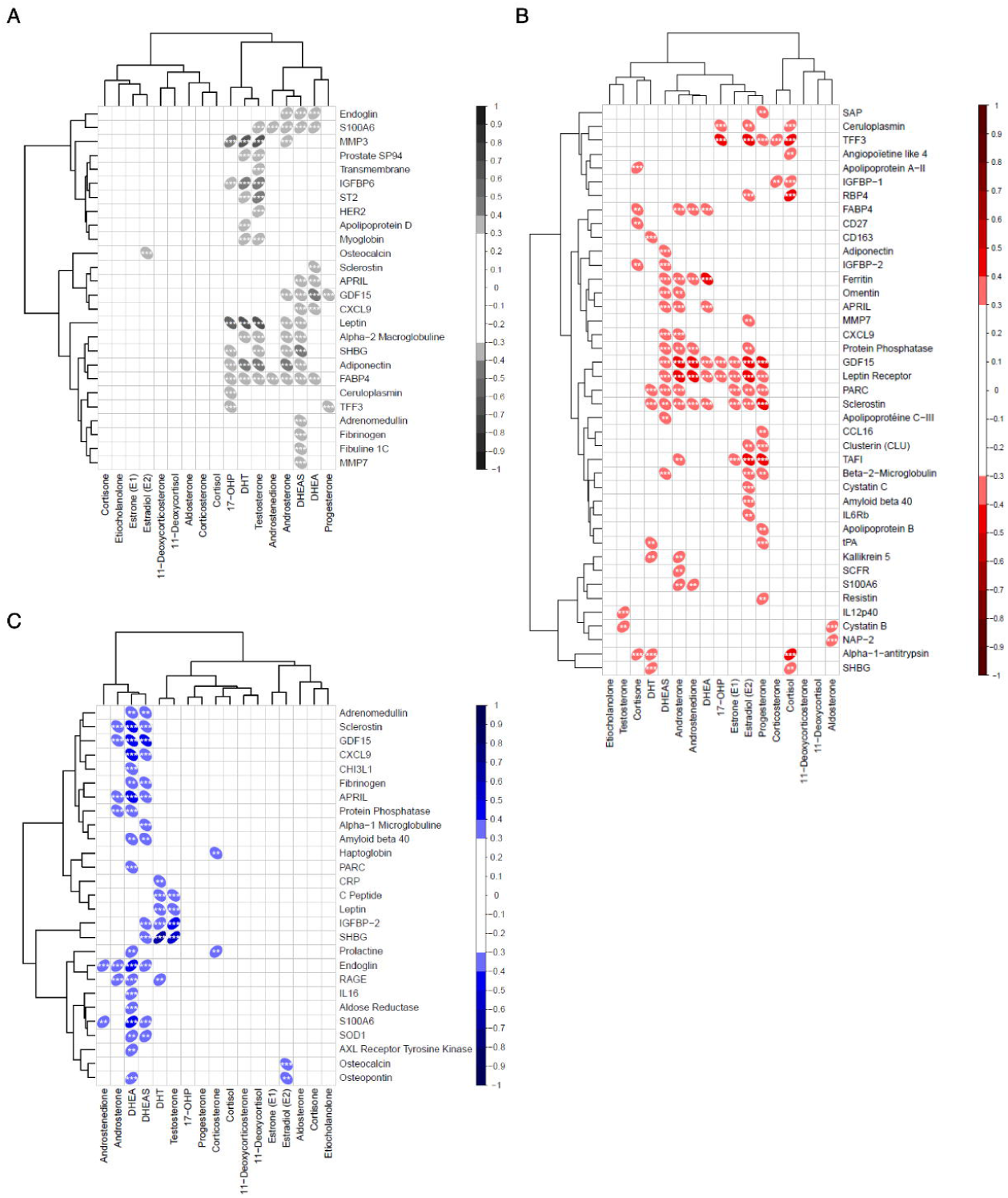
Plasma proteins have sex-specific associations with steroid hormone levels. **(A-C)** Heatmap of significant correlations between steroid hormone levels and 326 plasma soluble proteins from donors aged 30 to 39 and 60 to 69 with Spearman correlation R> |0.3| of **(A)** all donors **(B)** male donors and **(C)** female donors applying FDR correction (* = *q*<0.05, ** = *q*<0.01, *** = *q*<0.001). The direction of the oval indicates whether the relationship is positively or negatively correlated. (n=371 donors)

Several protein-steroid correlations were found only in the sex-specific analysis, primarily with androgens in male donors (**Figure 6B**). Interestingly the only correlation with E2 concentrations was for osteopontin and osteocalcin in male donors. Female-specific correlations were more numerous and observed with almost all the steroid hormones and a less defined clustering (**Figure 6B).** Interestingly, SHBG was not significantly correlated with DHT levels when analyzing all donors together, but was positively correlated with female and male donors when they were analyzed separately. Additionally, SHBG was negatively correlated with DHEA and positively correlated with testosterone only in male donors (**Figure 6B**) and positively correlated with cortisol in female donors (**Figure 6C**). Whether this finding is a sex specific mechanism, or due to limited statistical power remains unclear.

### Steroid hormone level variations are explained by sex, age, genetic, physiological variables and lifestyle

To determine the relative contributions of different factors explaining interindividual variability in steroid hormone levels, we performed a relative importance analysis, including sex, age, significant clinical variables (*q-*values > 0.01) and the lead causal SNPs (identified by fine mapping) for each hormone in the entire cohort. While sex explained a large part of the variance for 17-OHP, testosterone, and DHT, and age explained variance in DHEA, DHEAS, androstenedione and androsterone, much of the variance remained unexplained by the assessed parameters for the corticoids, estrogens, progesterone, and etiocholanolone (**Figure 7A**). Interestingly, compared to previously published studies for this cohort, in which sex explained on average 5% of variability in different immune phenotypes^21,32^, the impact of sex and age on steroids were major predictor variables. The same analysis was repeated independently for female and male donors to assess the sex-specific relative importance of the predictor variables (**Figure 7B-C**). We observed that while age still played an important role in sex-specific interindividual variability, oral contraceptive use had an impact on 10 of the 17 hormones measured (**Figure 7B**). The use and duration of concomitant drug treatments and menopausal status also explained some of the observed variance in female donors. In male donors, BMI and smoking status were the main predictor variables for explaining interindividual variability in 9 out of the 17 hormones (**Figure 7C**).

**Figure 7:**
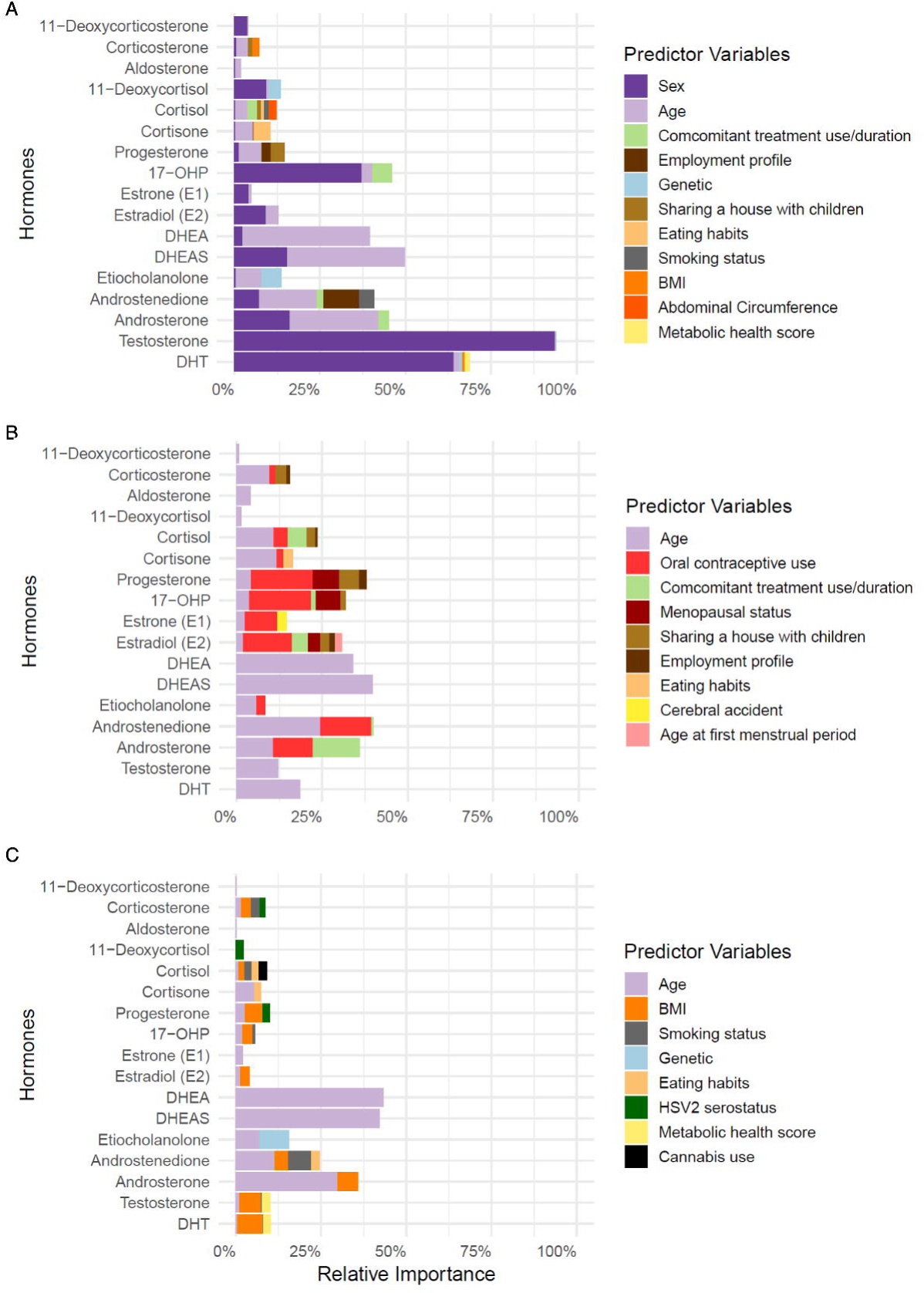
Sex and age explain a large proportion of interindividual variance in steroid hormone levels. **(A-C)** Graphs show the percentages of variance explained by each predictor for each hormone in **(A)** all donors n=949, **(B)** female donors n=472, or **(C)** male donors n=477. Average contribution of each predictor to the explained variance (R²) for each hormone is ordered by total importance across all hormones. Predictor variables with relative importance higher than 1% in at least one hormone are plotted.

### Steroid levels may reflect health and disease status

A feature of the original *Milieu Intérieur* study^22^ was its cross-sectional design with a narrow inclusion period. The inclusion of only healthy donors deliberately restricted the heterogeneity of the cohort, but in doing so limited extrapolation to disease development. To address this limitation, we recently re-recruited a subset (n=415) of the original cohort ten years after the initial study^33^, for a third sampling visit (V3). Importantly, all donors from the original study willing to participate in V3 were included, which allowed us to assess their health status 10 years after their original classification as healthy. MI V3 participants were assigned a status of “healthy”, “at risk”, or “disease” determined by whether they would meet the original inclusion/exclusion criteria (healthy), be excluded based on specific criteria (e.g., BMI) but were currently disease-free (at risk), or they reported a disease at the time of the V3 study (disease)^33^. Within these 415 donors, there was an age bias towards older age, but the equal sex balance was maintained^33^.

To test associations between changes in steroid hormone levels over the 10-year period with health status, we measured steroid hormones from this time point (V3) together with the samples measured in **Figure 1** (V1) in a randomized fashion to avoid technical batch effects. Due to the age bias in the disease group (**Figure 8A, B**), we corrected all measures by applying linear models with age, circular time, and circular date to correct for circadian and seasonal steroid cycles and used the residuals for subsequent analysis^34^. To study the relationship between steroid hormone changes over time and health status, we calculated the individual differences between V1 and V3 residual measures, hypothesizing that the disease group would show a greater decline over the 10-year period. Remarkably, we did not identify any significant hormone changes associated with health status in female donors (**Fig S7**). In male donors, we observed significant differences between healthy and disease groups for 11-deoxycorticosterone, progesterone, 17-OHP, and androstenedione **(Figure 8C)**. We plotted the regression of 11-deoxycorticosterone, progesterone, 17-OHP, and androstenedione separately for the three groups over the 10-year period, which revealed the steepest decline in these hormones in the disease group as reflected by their slopes **(Figure 8D)**. The healthy group had the smallest decline in every hormone except progesterone, where the at-risk group showed a slight increase **(Figure 8D)**.

**Figure 8:**
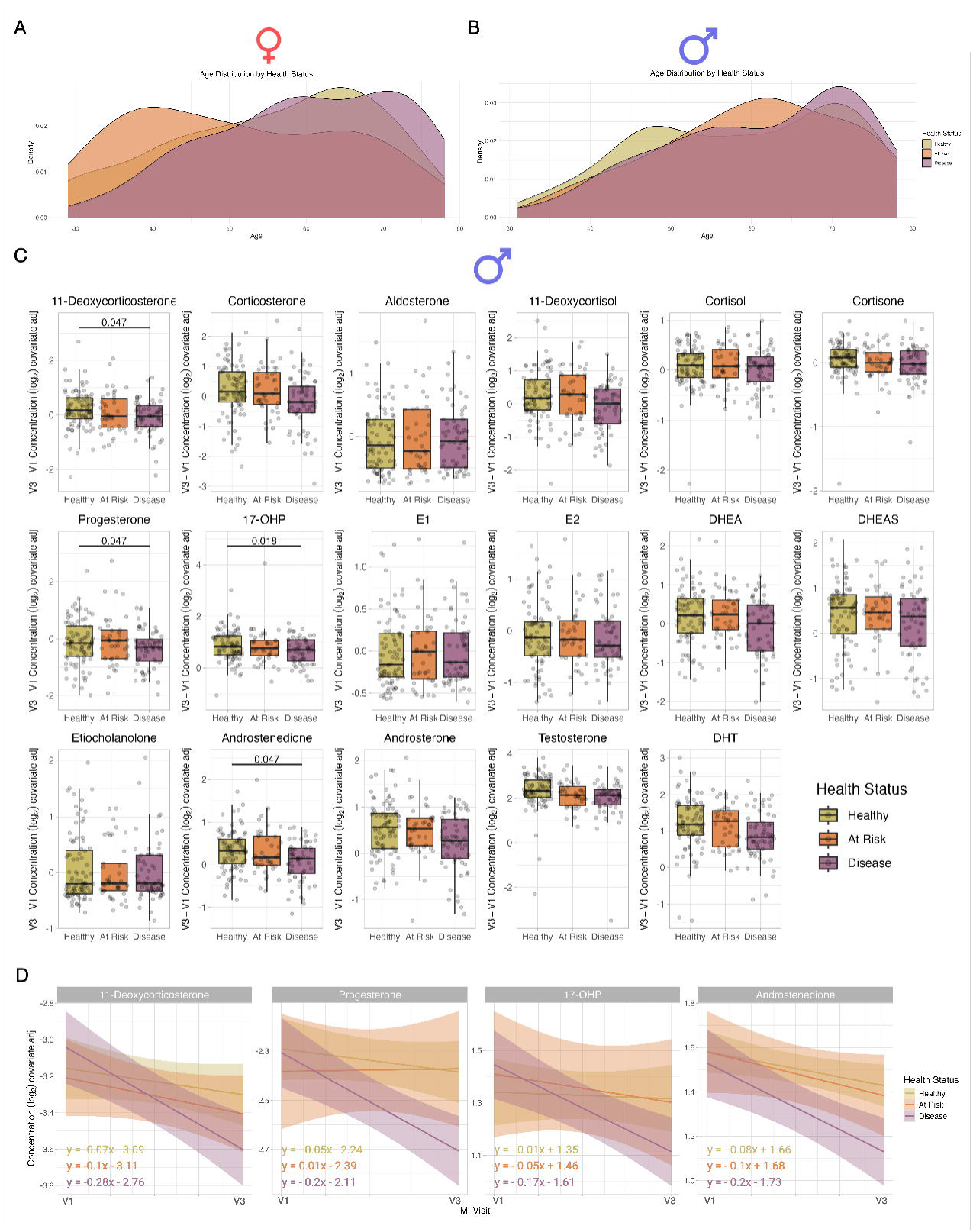
Steroid levels are significantly associated with a disease state only in men. (**A-B**) Ridge plot of the age distribution in each health group in (**A**) female donors and (**B**) male donors. (**C**) Box plots show the individual changes in covariate-corrected steroid hormone levels from V1 to V3 by health status in male donors. Significant *q*-values are displayed from Mann-Whitney U tests comparing at-risk and disease groups to the healthy group, with FDR correction across all tests. (**D**) Regressions of covariate-corrected steroid levels in male donors from V1 to V3 by health group for steroids with significant Mann-Whitney U results. (n=204 male donors)

## Discussion

This study describes diverse factors affecting steroid hormones in a well-defined healthy cohort balanced for sex and age, including a unique 10-year follow-up. Our analyses revealed expected findings, such as the decline of estrogens in older women compared to younger individuals. It also revealed unexpected findings such as the strong relationship between hormonal oral contraception and non-sex steroid hormones in women, and the wide-spread impact of BMI and smoking on steroid levels in men. It uncovered a relationship between specific steroid hormone decline over time and health status in men. Indeed, a strength of the original *Milieu Intérieur* cohort is the robust health of all donors due to the stringent inclusion and exclusion criteria. This allows the study of older individuals in good health and provided a homogenous initial health status for our comparisons in the 10-year follow-up study.

From this analysis, it suggests that the andropause and associated decline in testosterone in men may not be directly associated with aging, but rather with poor health. Another strength of our analyses is the ability to determine relationships between demographic data and steroid levels. Gender, economic and social status, and cultural habits influence behavior and lifestyle, with subsequent indirect effects on many biological processes^25^. We identified that many of these behaviors are significantly associated with one or more steroid hormone levels. Some associations, such as housing or employment, may be due to non-linear effects of age or other underlying factors, but are still important to consider in medical research.

Our study also has some important limitations. We have provided baseline sex and age-specific ranges for steroid hormone levels in a cohort that is genetically homogeneous (*i.e.*, European ancestry) and from a single geographical location. However additional large-scale studies are needed in individuals with diverse genetic ancestries and exposed to various environmental conditions^35^. A second limitation relates to sample size due to the large number of factors tested, and reduced statistical power with each subdivision of the cohort (*e.g.*, biological sex, menopausal, or health status) and when testing interaction effects. Because of this, possible biologically significant effects on steroids were not uncovered by our analyses.

Despite these limitations, our work raises new questions, such as the relationship between numerous steroid hormones and oral contraceptives, corticoids and activity levels or eating habits, or androgens and smoking behaviors in otherwise healthy individuals. Quite strikingly, although hormonal contraception is commonly used, as exemplified by 40% of pre-menopausal donors in our cohort, the impact of suppressing progesterone and estrogen over time is not well understood. We observed that hormonal contraception use was significantly associated with 12 out of 17 of the steroid hormones measured, and younger donors not using this form of birth control and menopausal women had non-sex steroid levels that were more similar to each other than to levels in women using oral contraception. Additionally, the levels of progestogens and estrogens in women using oral contraceptive pills were even lower than those found in postmenopausal women. The effects of this global alteration in steroid hormone levels in young women are unclear and raise the question as to whether broadly suppressing steroid hormones for prolonged periods of time impacts health status concurrently or later in life.

Many of the smoking variables were associated with androstenedione levels, which were specifically higher in current smokers compared to past or non-smokers. Interestingly, higher levels of androstenedione levels in smokers are described in different populations, although previous studies were performed separately in women^36–38^ or men^39^. While we only found this significant association in male donors, the proportion of female smokers was similar (19% and 23%, respectively). Alternatively this lack of statistical significance could reflect other complex confounding factors, for example male smokers may be heavier smokers and are significantly more likely to be of low SES compared to nonsmokers, which is not the cases for female smokers^25^. Smoking status was also associated with E2 levels in women, which is in line with a report that serum E1, E2, and estriol (E3) concentrations are lower in active and passive female smokers compared to nonsmokers^40^. These differences were uncovered when we analyzed female and male donors separately, underlining the importance of studying both sex and gender effects. We recently reported that smoking has reversible impacts on innate immune parameters, but long-lasting effects on adaptive immunity^32^. Whether these impacts on immunity may be linked to effects on steroid hormone levels remains to be determined, however smoking habits should be considered when treating hormone-associated diseases or using hormone modifying treatments.

All steroid hormones showed a significant association with one or more plasma proteins measured, while 6 had significant genetic associations. These analyses are likely impacted by the complex regulation of free and bound steroids in circulation supported by the strong associations revealed with SHBG, in both genetic and plasma protein analyses. Associations between etiocholanolone and variants upstream of UGT2B17, an enzyme involved in the glucuronidation of certain androgens, may be due to metabolic elimination of this steroid. The addition of glucuronic acid from UDP-glucuronic acid to etiocholanolone leads to increased water solubility and facilitates excretion in urine^41^.

In summary, we present new insights into the causes of variability in steroid hormones in healthy women and men aged 20-79 years old. Overall, sex, age, genetics, plasma protein levels, and lifestyle choices influence steroid hormone levels, however, how these complex relationships are mediated remains to be determined. These hypothesis-generating findings will support future mechanistic studies of steroid hormone levels and key immune functions in non-infectious, hormone exacerbated diseases. It may serve as an important resource that will benefit the research community, not only in endocrinology and endocrine system-associated diseases, but all fields in which steroid hormones play a role, including metabolism, immunology, and aging.

## Materials and Methods

### Clinical Protocol

Human samples were from the *Milieu Intérieur* cohort, which was approved by the *Comité de Protection des Personnes*-Ouest 6 on June 13th, 2012, and by the French *Agence Nationale de Sécurité du Médicament* (ANSM) on June 22nd, 2012. The study was sponsored by Institut Pasteur (Pasteur ID-RCB Number: 2012-A00238-35) and conducted as a single center interventional study without an investigational product. The original protocol was registered under ClinicalTrials.gov (study# NCT01699893). The samples and data used in this study were formally established as the *Milieu Intérieur* biocollection (NCT03905993), with approvals by the *Comité de Protection des Personnes - Sud Méditerranée* and the *Commission Nationale de l’Informatique et des Libertés* (CNIL) on April 11, 2018. Donors gave written informed consent. The 1,000 donors of the *Milieu Intérieur* cohort were recruited by BioTrial (Rennes, France) and were composed of “healthy” individuals of the same genetic background (Western European) and to have 100 female donors and 100 male donors from each decade of life, between 20 and 69 years of age. Donors were selected based on various inclusion and exclusion criteria that were previously described and self-declared their gender^22^. To avoid the influence of hormonal fluctuations in female donors, pregnant and peri-menopausal women were not included. The recruitment of donors was restricted to individuals whose parents and grandparents were born in metropolitan France and that had no family relationships, which minimizes genetic stratification.

The 10 year follow up “V3” study was approved by the *Comité de Protection des Personnes -Nord Ouest III* (Committee for the protection of persons) on 27th January 2022, and by the French Agence nationale de sécurité du médicament (ANSM) on 30th November 2011^33^. The study is sponsored by the Institut Pasteur and was conducted as a single center study without any investigational product. The protocol is registered at ClinicalTrials.gov (study# NCT05381857). As this study was a 10 year follow up study of the original Milieu Interieur “V1” study, the major inclusion criterion was previous inclusion in these studies. A health score labelled each V3 participant as “healthy”, “at risk”, or “disease” depending on the original MI inclusion criteria: whether they would still be included, they would not pass inclusion, or they have a reported diagnosis, respectively. In addition, donors were required to give written informed consent and be affiliated to the French social security regimen.

### Steroid hormone measurements

For free steroid hormone measures plasma samples from fasting EDTA-treated blood was frozen immediately upon collection at -80°C until aliquoting. Upon thawing, aliquoted samples were completely randomized prior to processing for mass spectrometry analysis. 17 steroid hormones were measured using the AbsoluteIDQ Stero17 kit (Biocrates, Austria), which included glucocorticoids (cortisol, cortisone, and 11-deoxycortisol), mineralocorticoids (aldosterone, corticosterone, and deoxycorticosterone [DOC]), progestogens (17α-hydroxyprogesterone [17-OHP] and progesterone), estrogens (estrone [E1] and estradiol [E2]), and androgens (androstenedione, androsterone, dehydroepiandrosterone [DHEA], dehydroepiandrosterone sulfate [DHEAS], dihydrotestosterone [DHT], etiocholanolone, and testosterone). Measurements of the 17 hormones were obtained from Biocrates in an Excel sheet along with a corresponding data report. Given that the measurements were taken across multiple plates, Biocrates calculated a project-based limit of detection (LOD), corresponding with the highest LOD in any plate for each steroid hormone. For measurements below LOD, Biocrates imputed missing values using the R package logspline. The logspline imputation method generates values between LOD and LOD/2, and applies the variance between values above LOD. Therefore, if there is high variance in the dataset, the imputed values may also exceed these limits.

### Datasets

*Case Report Form variables:* Lifestyle, dietary, and medical history factors were collected in an electronic case report form (CRF)-based questionnaire as previously described^22^. To select factors for analysis, we chose those variables that had at least 5% of individuals in at least two categorical levels. The metabolic score, which varies from 0 to 1, was previously computed for each donor^22^ by incrementing the score by 1 when: abdominal circumference >94cm and >80cm in male or female donors, respectively; systolic blood pressure ≥ 130mmHg or diastolic blood pressure ≥ 85mmHg; triglyceride levels ≥1.7mM; HDL levels < 1 mM and < 1.3 mM in male or female donorss, respectively; glucose concentration ≥ 6.1 mM ^23^.

### Genetic data

Protocols and quality control filters for genome-wide SNP genotyping were previously described^21^. Briefly, the 1,000 *Milieu Intérieur* donors were genotyped on both the HumanOmniExpress-24 and the HumanExome-12 BeadChips (Illumina, California, USA), which include 719,665 SNPs and 245,766 exonic SNPs, respectively. Variants that were unmapped on dbSNP138, duplicated, with low genotype clustering quality (GenTrain score < 0.35), with a call rate of < 99%, or monomorphic were filtered out. For the X chromosome, SNPs that were heterozygous in >1% of males were removed. SNPs were further filtered for minor allele frequencies > 5%, yielding a data set composed of 1,000 donors and 5,715,171 SNPs for analysis.

### Plasma protein measures

Plasma proteins were previously described^31^. Briefly 326 soluble blood proteins were measured by multi-analyte Luminex assays (Rules Based Medicine) in the plasma of a subset of 400 donors.

### Data analysis

Unless otherwise stated, all displayed results were performed on the 949 individuals of the cohort who gave consent to share their data publicly. For analysis, the steroid hormones measures were log_2_ transformed. Samples were prepared for shipments in two batches. Possible aliquoting batch effects were assessed using linear modelling. Batch effects were corrected using the removebatcheffect function from Limma in R (version 4.2.2), where age and sex were included in the design parameter as biological variables that should be maintained. PCA plots were produced with Qlucore Omics Explore (V3.9.9).

For visualization of steroid levels with age and sex, a loess-function was applied with a span of 0.75 and degree of 1. A linear model with the covariates of age, sex, and their interaction was applied to the steroid hormone levels. The subsequent estimate, minimum, and maximum were extracted from the models to calculate the effect sizes of these covariates. An age-sex interaction term was incorporated in all subsequent models.

### eCRF analysis

To test for associations between demographic variables and steroid hormone levels we used likelihood ratio tests comparing a base model controlling for age, sex and an age×sex interaction [lm(hormone ∼ age×sex)], with a model controlling for age, sex and an age sex interaction plus a predictor demographic variable [lm(hormone ∼ age×sex + demographic variable)]. We used chi-square tests for the LRT. We performed this analysis on 137 demographic variables in all donors, with an additional 14 variables in female only donors. An FDR correction was performed to account for multiple testing across all demographic features for each hormone. Heatmaps to display the q value from the LRT were generated using the R package pheatmap. For the residual plots, *q-*values were calculated using a Kruskal Wallis with FDR correction for all tests together followed by post-hoc Dunn’s test.

### Genetic association analysis

Genome wide association testing was performed for each hormone using PLINK (version 1.9). Analysis was performed in males and females separately and the results of both sexes were combined using meta-analysis. Covariates included for both sexes were age, BMI, smoking status, as well as PC1 and PC2 of the genetic PCA to account for potential population structure in the study-cohort^42^. Additional covariates included in the female specific analysis included menopausal status, HRT, oral contraceptive use, IUD, and tubal ligation. Meta-analysis was used to combine the results of male and female donors for each hormone using PLINK (version 1.9). A *p*-value of *p*<5x10^-8^ was taken as significant. QQ plots with λ values were used for quality control. Locus zoom plots were generated using the R package locuszoomr. Linkage disequilibrium for the plots was determined using data from the 1000 genomes project using the lead SNP identified by fine mapping using susieR as the reference. Manhattan plots were generated using ggplot2. Data for residual plots of individual SNPs were generated by running a linear model including all of the GWAS covariates using the tidy and augment functions from the R package broom. Male donors were considered as answering “no” for the female only covariates for the purposes of this adjustment. Plots were generated using the ggpubr package.

### Fine mapping of genome wide significant SNPs

Statistical fine mapping was performed on meta-analysis data using the susieR R package (version 0.12.35) in a 1Mb window centered on the midpoint of significant SNPs within in the same locus (https://doi.org/10.1111/rssb.12388). Z scores were generated from summary statistics by dividing beta values by the standard error. LD matrices of r values were generated using PLINK v1.9 using Milieu Interieur data. The susie_rss function was used with the maximum number of credible sets (L) set to 10, and the number of iterations set to 100. For each locus, the SNP with the highest posterior inclusion probability (α) was selected as the index SNP.

### Longitudinal data

For comparison of steroid hormones over the 10-year sampling period, the log_2_-transformed concentrations were adjusted for age as well as circadian and annual rhythms by applying a linear model with age, and the sines and cosines of day of the year and time of day as the only covariates and extracting the subsequent residuals. The difference in adjusted steroid hormone concentrations over 10 years was calculated for each donor, then divided by sex and grouped by health score. The adjusted hormone differences of the at risk and disease groups tested against the reference healthy group via Mann-Whitney U, and subsequent p-values were adjusted for multiple testing with FDR correction. An FDR q-value less than or equal to 0.05 was considered significant.

### Computation of variance explained

For each steroid hormone, sex, age, the most significant clinical variables (q values > 0.01), and the causal SNPs identified by fine mapping were considered to calculate the percentage of variance explained by each of these predictor variables with R package relaimpo 2.2.7 and plotted with the R package ggplot2 3.5.1. The R^2^ contribution averaged over orderings among variables was computed using the lmg type in the calc.relimp function of the relaimpo R package. For each hormone, genetic (rs34253002 for etiocholanolone, rs149932962 for testosterone and DHT and rs4523233 for 11-deoxycortisol in all donors and rs138863991 for etiocholanolone in male donors), clinical variables such as eating habits (eating only at night or during the evening, eating fast food and eating lunch), employment profile (employment status, working time per day and steady job) and all the other most significant clinical variables (q values > 0.01) per hormone were considered in the model.

### Data availability

SNP array data can be accessed from the European Genome-Phenome Archive EGA under accession EGAS00001002460. The steroid hormone data set can be accessed by requesting to.

## Data Availability

All data produced in the present study are available upon reasonable request to the authors, or are available online.

https://ega-archive.org/studies/EGAS00001002460

https://dataset.owey.io/doi

## Acknowledgements

The Milieu Intérieur Consortium¶ is composed of the following team leaders: Laurent Abel (Hôpital Necker), Andres Alcover, Hugues Aschard, Philippe Bousso, Nollaig Bourke (Trinity College Dublin), Petter Brodin (Karolinska Institutet), Pierre Bruhns, Nadine Cerf-Bensussan (INSERM UMR 1163 – Institut Imagine), Ana Cumano, Christophe D’Enfert, Ludovic Deriano, Marie-Agnès Dillies, James Di Santo, Gérard Eberl, Jost Enninga, Jacques Fellay (EPFL, Lausanne), Ivo Gomperts-Boneca, Milena Hasan, Gunilla Karlsson Hedestam (Karolinska Institutet), Serge Hercberg (Université Paris 13), Molly A Ingersoll (Institut Cochin and Institut Pasteur), Olivier Lantz (Institut Curie), Rose Anne Kenny (Trinity College Dublin), Mickaël Ménager (INSERM UMR 1163 – Institut Imagine), Frédérique Michel, Hugo Mouquet, Cliona O’Farrelly (Trinity College Dublin), Etienne Patin, Antonio Rausell (INSERM UMR 1163 – Institut Imagine), Frédéric Rieux-Laucat (INSERM UMR 1163 – Institut Imagine), Lars Rogge, Magnus Fontes (Institut Roche), Anavaj Sakuntabhai, Olivier Schwartz, Benno Schwikowski, Spencer Shorte, Frédéric Tangy, Antoine Toubert (Hôpital Saint-Louis), Mathilde Touvier (Université Paris 13), Marie-Noëlle Ungeheuer, Christophe Zimmer, Matthew L. Albert (Octant Biosciences), Darragh Duffy§, Lluis Quintana-Murci§,

¶ unless otherwise indicated, partners are located at Institut Pasteur, Paris

§ co-coordinators of the Milieu Intérieur Consortium

Additional information can be found at: http://www.milieuinterieur.fr

Figures were compiled using BioRender.

## Author contributions

MAI, DD conceived, developed, and supervised the study. LD, JS, EM performed data analysis. LD, JS, EM, MAI, DD wrote the manuscript. FD prepared and managed human samples. AJ, JB, EP provided data management and analytical support. LQM, MAI, DD obtained funding and coordinated the clinical samples. All authors reviewed the manuscript prior to submission.

## Data and materials availability

Further information and requests for resources should be directed to and will be fulfilled by Molly A. Ingersoll and Darragh Duffy. Requests for Milieu Interieur data sets should be sent to.

## List of Supplementary data

**Supplementary Figure 1:** Steroid hormones are interconnected in biochemical steroidogenesis pathways.

**Supplementary Figure 2:** Steroid hormone level correlations separately for female and male donors.

**Supplementary Figure 3:** Relative steroid hormone levels among a cohort of healthy donors.

**Supplementary Figure 4:** Steroid hormone SNP plots for male donors.

**Supplementary Figure 5:** Steroid hormone SNP plots for all donors.

**Supplementary Figure 6:** Steroid hormone SNP plots for all donors.

**Supplementary Figure 7:** Steroid levels are not significantly associated with disease state in female donors.

