## Supplemental Figures 1-7 for "Steroid hormone levels vary with sex, aging, lifestyle, and genetics"

Supplementary Figures

### Supplementary Figure 1

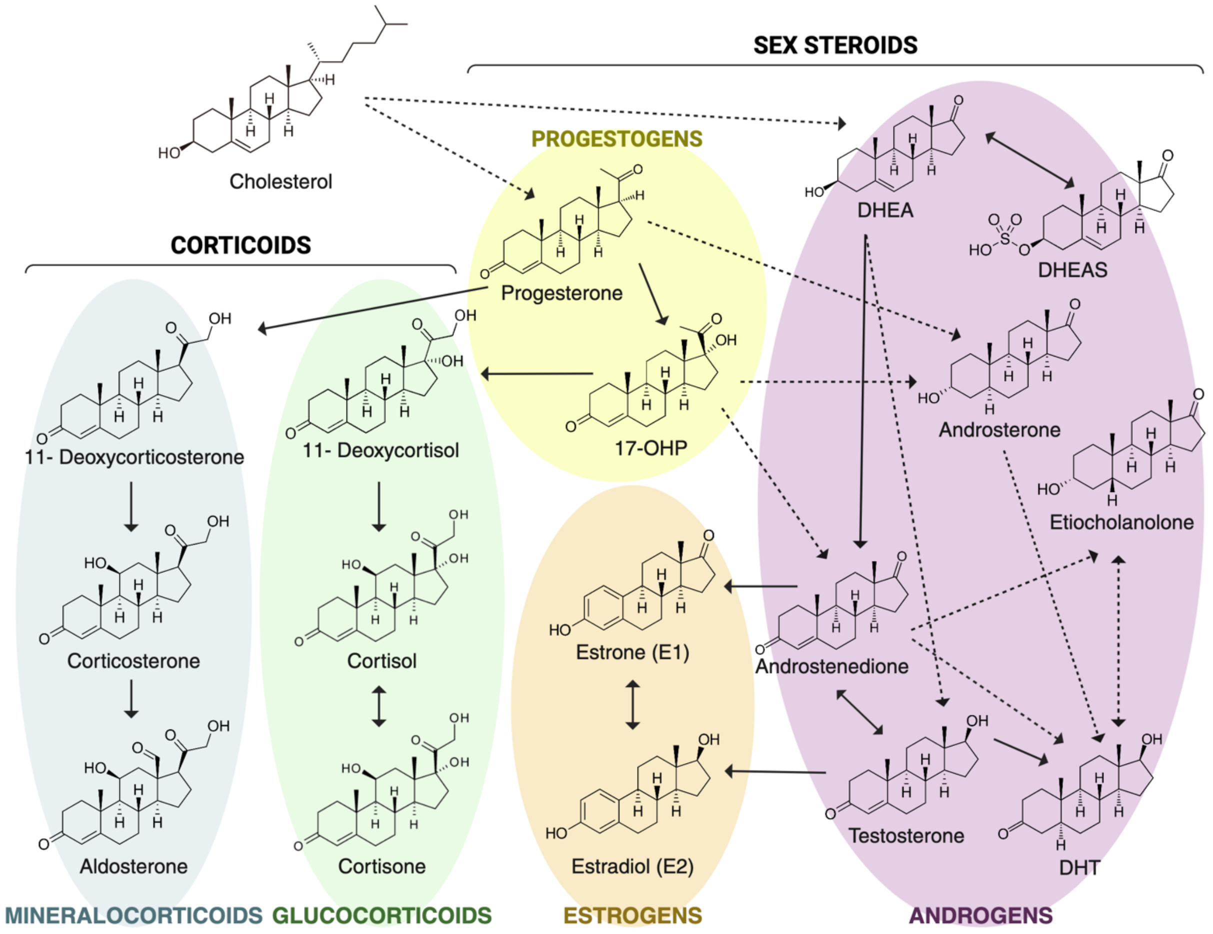


**Supplementary Figure 1: Steroid hormones are interconnected in biochemical steroidogenesis pathways.** Schematic overview of steroidogenesis and the relationship among the steroid hormones measured in this study. Steroids are grouped into 2 categories: corticoids and sex steroids, which are then separated into 5 groups: mineralocorticoids, glucocorticoids, progestogens, estrogens, and androgens. Arrows show direct relationships between steroids, such as conversion by one enzyme. Dotted line arrows show relationships with at least one intermediate and two enzymes between the two steroids shown. 17-OHP: 17-hydroxyprogesterone, DHT: dihydrotestosterone, DHEA: dehydroepiandrosterone, DHEAS: dehydroepiandrosterone sulfate. Adapted from [1,41,43].

**Supplementary Figure 2**

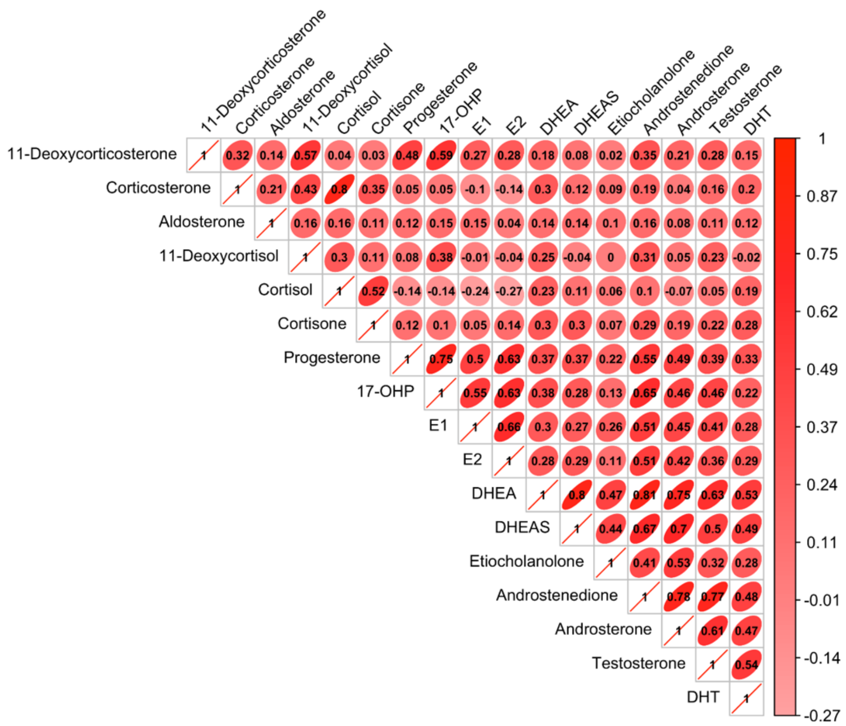


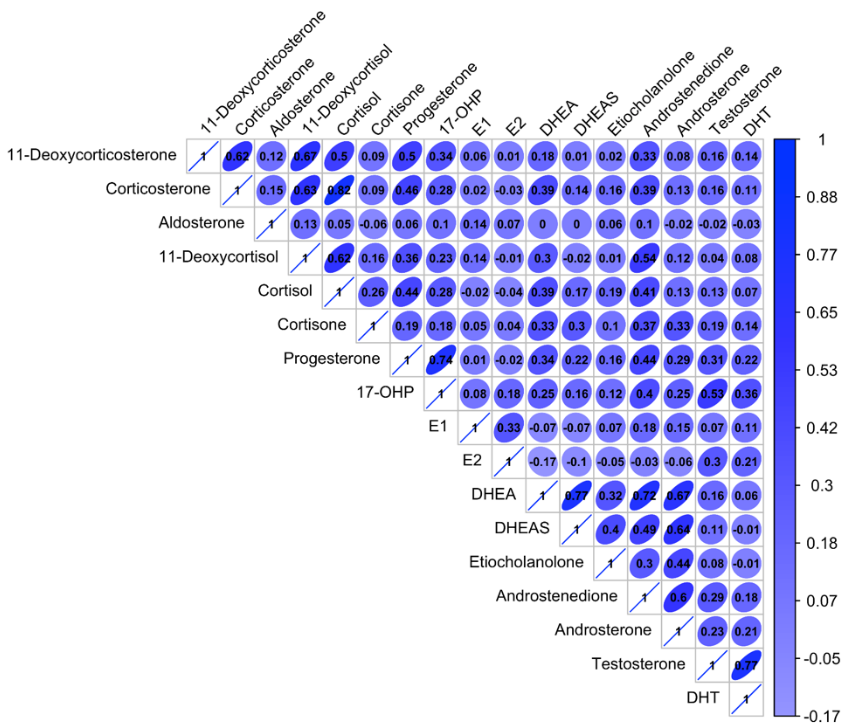


**Supplementary Figure 2: Correlations between steroid hormone levels separately for female and male donors.** Correlation matrices showing Spearman correlation between hormones using $log_2$ data without adjusting for age separately for female (red) and male (blue) donors. $r^2$ values are displayed on the plot. Ellipses indicate the direction of correlation. n=472 females, n=477 males.

**Supplementary Figure 3**

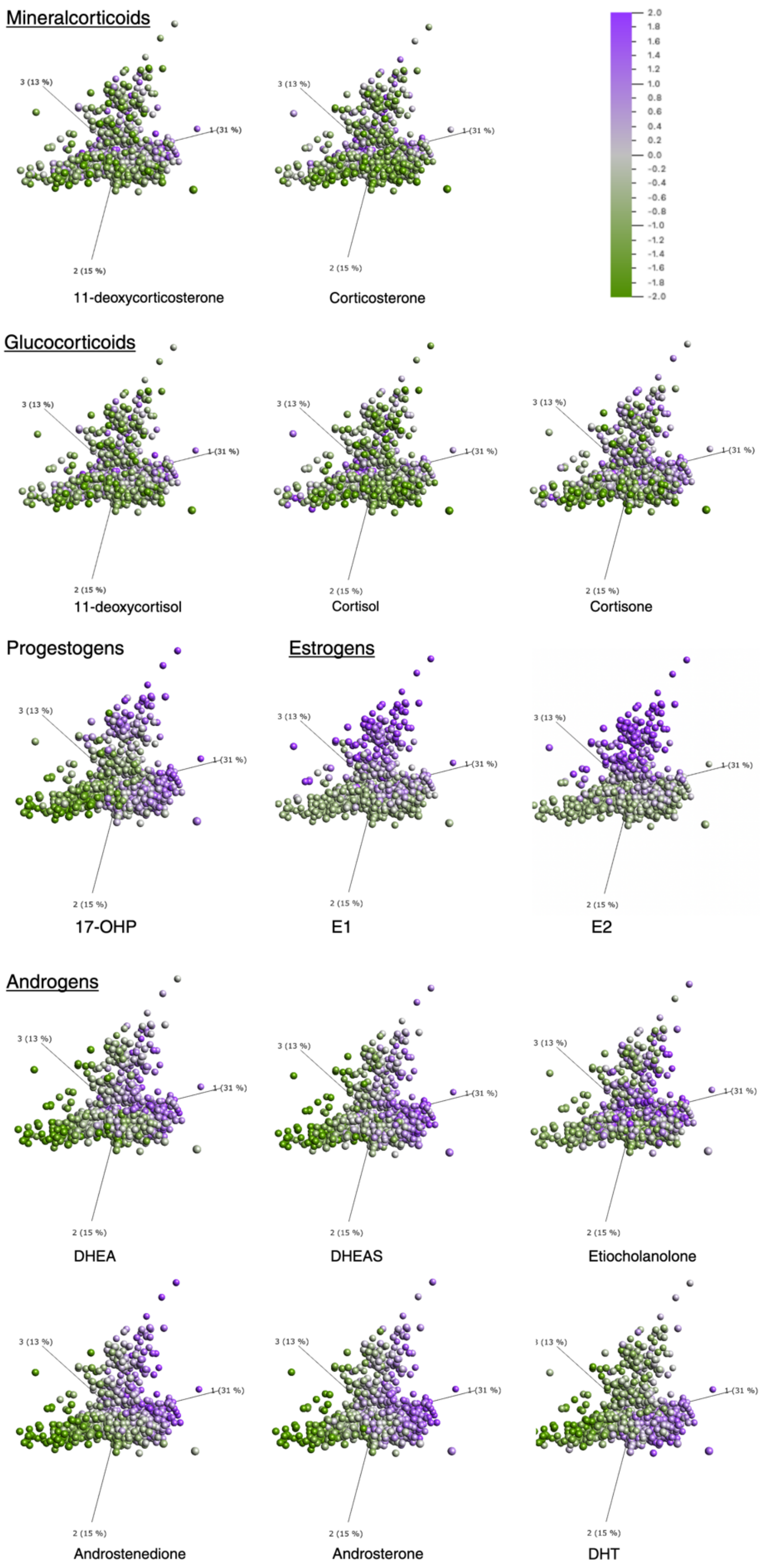


**Supplementary Figure 3: Relative steroid hormone levels among a cohort of healthy donors.** PCA of log-transformed nanomolar concentrations of steroid hormones measured by LC-MS/MS color coded by relative expression levels per hormone. (n=949 donors)

**Supplementary Figure 4:**

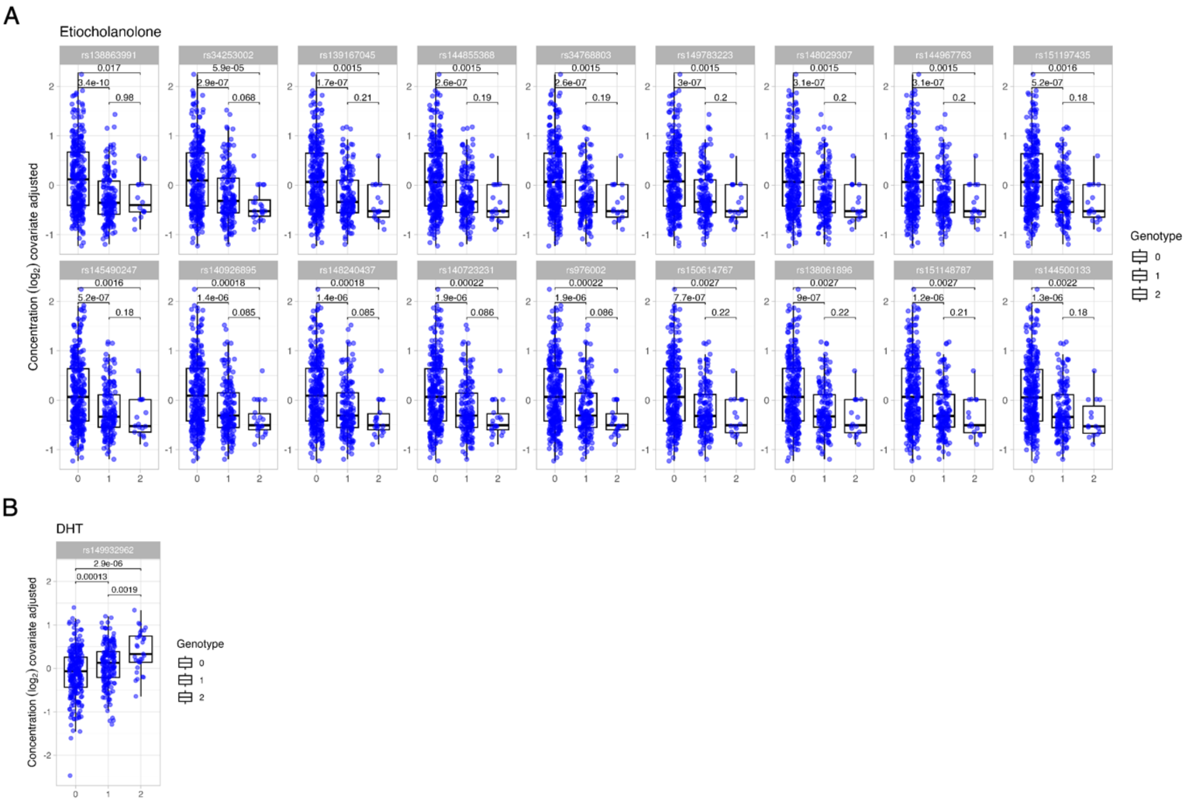


**Supplementary Figure 4: SNPs associated with etiocholanolone and 11-deoxycortisol levels in male donors.** Box plots of (A) etiocholanolone and (B) DHT levels in male donors for significant SNPs identified in the GWAS. Data has been corrected for covariates included in the GWAS (age, BMI, smoking status, as well as PC1 and PC2 of a genetic PCA, menopausal status, HRT, oral contraceptive use, IUD, and tubal ligation). P values displayed here are from a Dunn's test with an FDR adjustment. (n=477)

### Supplementary Figure 5:

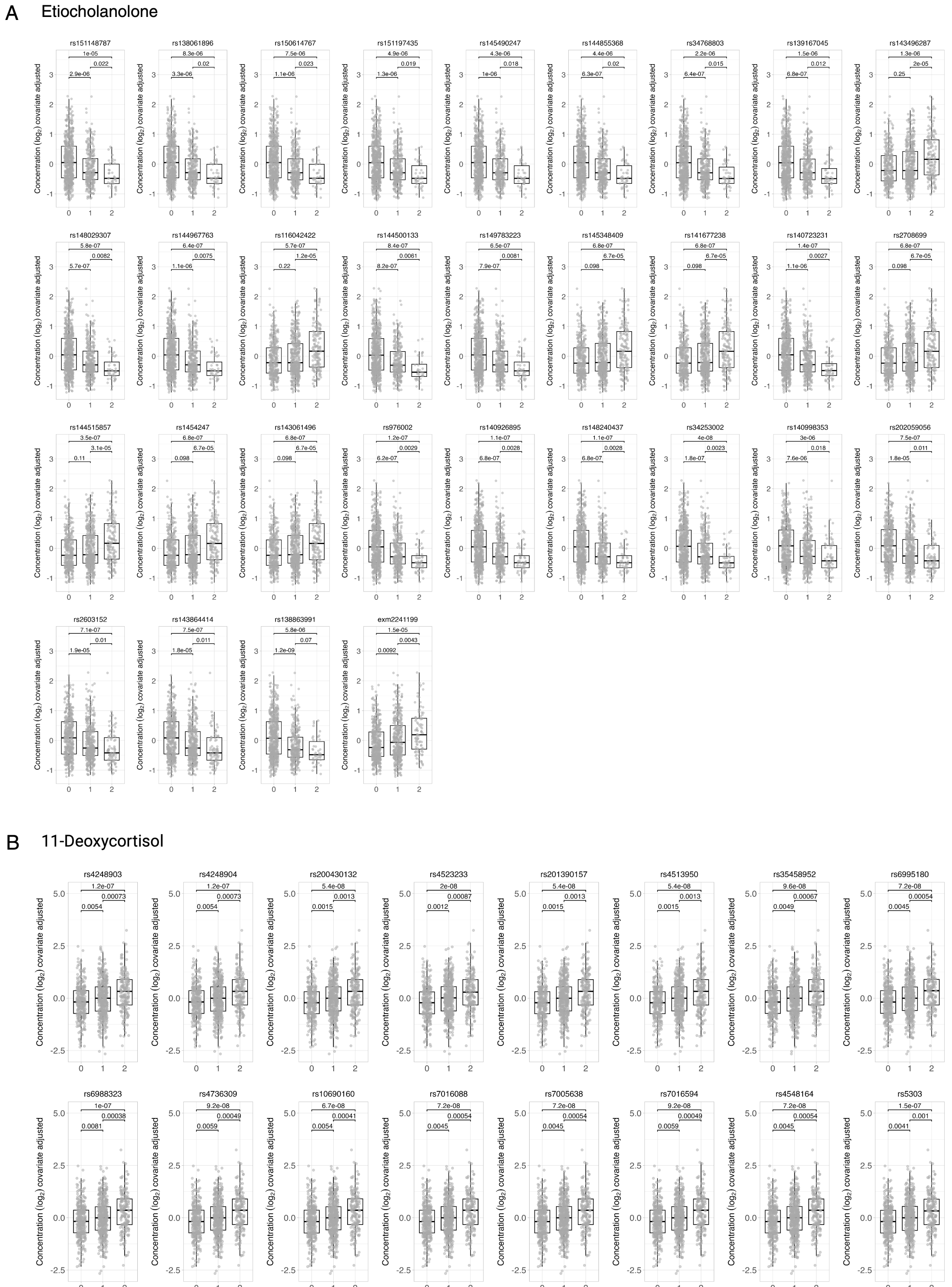


**Supplementary Figure 5: SNPs associated with etiocholanolone and 11-deoxycortisol levels in all donors.** Box plots of (**A**) etiocholanolone and (**B**) 11-deoxycortisol levels in all donors for significant SNPs identified in the GWAS. Data has been corrected for covariates included in the GWAS (age, BMI, smoking status, as well as PC1 and PC2 of a genetic PCA, menopausal status, HRT, oral contraceptive use, IUD, and tubal ligation). P values displayed here are from a Dunn's test with an FDR adjustment. (n=949 donors)

**Supplementary Figure 6:**

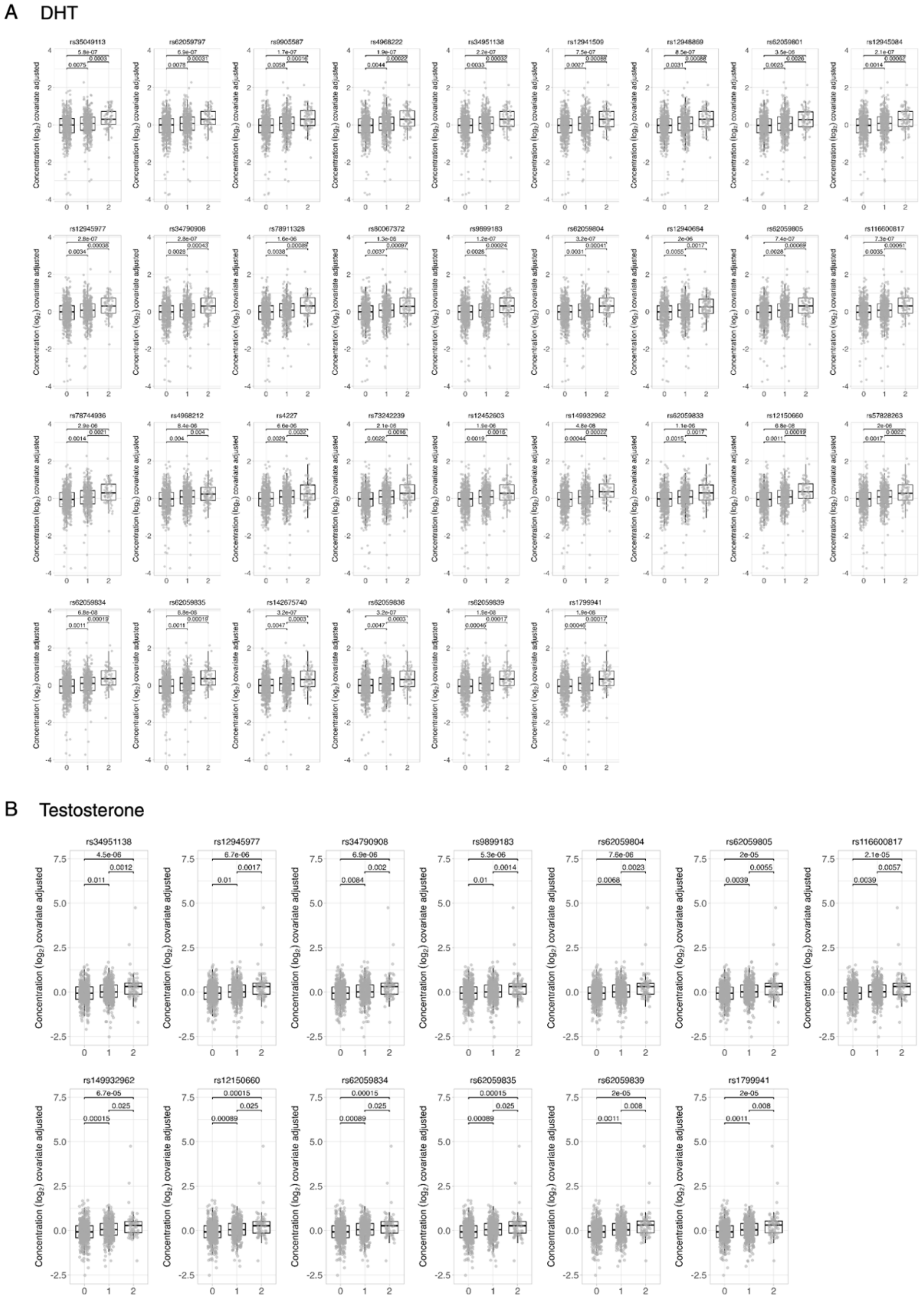


**Supplementary Figure 6: SNPs associated with DHT and testosterone levels in all donors.** Box plots of (A) DHT and (B) testosterone levels in all donors for significant SNPs identified in the GWAS. Data has been corrected for covariates included in the GWAS (age, BMI, smoking status, as well as PC1 and PC2 of a genetic PCA, menopausal status, HRT, oral contraceptive use, IUD, and tubal ligation). P values displayed here are from a Dunn's test with an FDR adjustment. (n=949 donors)

**Supplementary Figure 7**

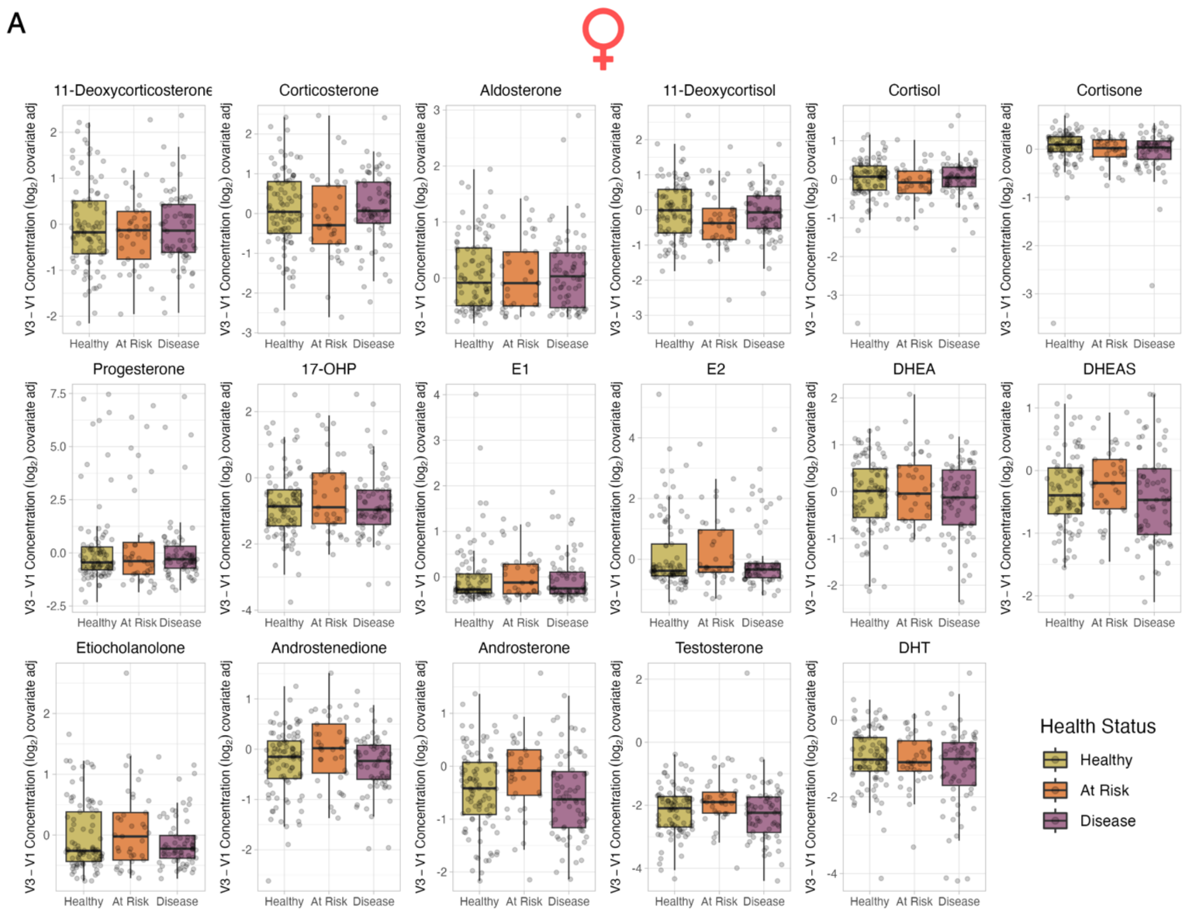


**Supplementary Figure 7: Steroid levels are not significantly associated with disease state in female donors.** (**A**) Box plots show the individual changes in covariate-corrected steroid hormone levels from V1 to V3 by health status in female donors. There were no significant *q*-values from Mann-Whitney U tests comparing at-risk and disease groups to the healthy group, with correction across all tests. (n=209)
